# Context-dependent molecular responses to heterogeneous metabolic disease traits

**DOI:** 10.64898/2026.05.31.26354544

**Authors:** Theodora-Dafni Michalettou, Sapna Sharma, Natalie N. Atabaki, Juan J. Fernandez-Tajes, Mun-gwan Hong, Jonathan Adam, Jerzy Adamski, Søren Brunak, Federico De Masi, Emmanouil T. Dermitzakis, Ian M. Forgie, Paul W. Franks, Tim Frayling, Giuseppe N. Giordano, Mark Haid, Torben Hansen, Tue H. Hansen, Peter P. Harms, Andrew T. Hattersley, Ulrik Plesner Jacobsen, Angus G. Jones, Robert W. Koivula, Tarja Kokkola, Markku Laakso, Anubha Mahajan, Andrea Mari, Mark I McCarthy, Timothy J. McDonald, Imre Pavo, Oluf Pedersen, Anna Ramisch, Hartmut Ruetten, Femke Rutters, Jochen M. Schwenk, Leen ’t Hart, Henrik Vestergaard, Mark Walker, Ewan R. Pearson, The IMI DIRECT Consortium, Andrew A. Brown, Ana Viñuela

## Abstract

Metabolic diseases such as type 2 diabetes (T2D) arise through complex interactions between physiological, molecular, and environmental processes. However, molecular studies often examine traits such as age, sex, adiposity, and glycaemic status independently, overlooking how their combined effects shape molecular phenotypes and relationships across molecular layers. Here, integrating transcriptomic, proteomic, metabolomic, and genetic data from 3,027 individuals in the IMI DIRECT cohort, we investigated the joint effects of age, sex, BMI, and HbA1c on molecular phenotypes and cross-omic relationships. We identified widespread molecular associations alongside extensive context-dependent effects, demonstrating that the effect of one trait frequently depended on the state of another, with sex-dependent age effects particularly prominent. These interactions were also evident in cross-omic relationships, indicating that physiological and metabolic context shapes not only molecular abundance but also the coordination between transcripts, proteins, and metabolites. Probabilistic causal inference further identified directionally structured relationships across molecular layers, with age-associated effects more frequently supporting dependent models consistent with inter-omic mediation. Together, our findings demonstrate that metabolic disease-relevant traits jointly shape molecular phenotypes and cross-omic network organisation. This context dependence highlights the importance of accounting for physiological state when interpreting molecular mechanisms and biomarkers, and provides a framework for investigating dynamic molecular signatures that may inform future approaches to disease stratification and precision medicine.

## INTRODUCTION

Metabolic diseases, and in particular type 2 diabetes (T2D), are complex systemic disorders arising from the interaction of multiple physiological, molecular and environmental processes. Clinical traits including body mass index (BMI), age, glycaemic status and sex each contribute to disease risk through distinct yet interconnected mechanisms. For example, increased adiposity, particularly visceral fat accumulation, promotes insulin resistance and chronic low-grade inflammation^1,2^, while ageing is associated with progressive metabolic stress, altered mitochondrial function and increased susceptibility to metabolic dysfunction^3–5^. Glycated haemoglobin (HbA1c), a marker commonly used for the diagnosis and prognosis of T2D which reflects long-term glycaemic exposure, captures cumulative disturbances in glucose homeostasis and disease progression^6,7^. HbA1c levels have also been positively correlated with age irrespective of T2D status^8,9^. Sex further influences metabolic disease trajectories, with men typically developing T2D at lower BMI thresholds, whereas women often exhibit delayed onset but greater cardio-metabolic complications following menopause^10,11^. Importantly, these traits do not act in isolation: their interactions contribute to the substantial clinical and biological heterogeneity of T2D that can manifest through differing contributions from adiposity, insulin resistance and pancreatic β-cell dysfunction^12–14^. For example, individuals included in the mild age-related diabetes (MARD) T2D subtype show an older age at onset relative to other subtypes; while patients in the severe insulin-resistant diabetes (SIRD) subtype show a low HbA1c and a high BMI^15^. Genetic studies of T2D subtypes and disease heterogeneity have identified different genetic architectures^16–19^, suggesting genetic regulation of the physiological and clinical variables may contribute to disease heterogeneity.

Understanding T2D heterogeneity requires investigating not only how physiological traits are associated with individual molecular phenotypes, but also how they shape the relationships between molecules. Metabolic processes depend on coordinated interactions across molecular layers, including gene expression, protein abundance and metabolite levels, which reflect the integration of biological processes rather than changes in individual molecules alone. However, many molecular studies focus on associations between clinical traits and individual molecular phenotypes, often implicitly assuming that relationships between molecules are broadly stable across individuals and physiological states. This assumption may obscure important context-dependent features of metabolic disease biology, in which the same molecular phenotype may participate in different relationships depending on an individual’s age, sex, adiposity or glycaemic status. This is particularly relevant given the dynamic nature of metabolic systems and the substantial evidence that molecular phenotypes vary with physiological context. BMI, a major risk factor for T2D^20–22^, is strongly associated with widespread changes in molecular phenotypes^23–28^ and typically increases with age, contributing to age-dependent variation in metabolic risk^29–31^. Ageing and obesity share biological hallmarks of metabolic and cellular dysfunction^32^. In parallel, age is associated with broad changes across the human transcriptome, proteome and metabolome^33–38^, while sex-specific differences in gene expression have been widely reported^39^, including divergent regulation of mitochondrial and immune pathways during ageing^40–42^. Together, these findings suggest that physiological traits may influence not only molecular abundance but also the coordination between molecular processes. However, whether and how age, sex, BMI and glycaemic status modify relationships between molecular phenotypes across omics layers remains insufficiently characterised. Analyses that assume stable cross-omic relationships may therefore overlook biologically relevant differences in the organisation of metabolic processes across physiological contexts.

Multi-omics approaches provide an opportunity to address this question by examining molecular phenotypes and their relationships across transcriptomic, proteomic and metabolomic layers. Previous studies integrating these data types have identified coordinated molecular changes associated with obesity, insulin resistance and T2D^43,44^, demonstrating the value of investigating metabolic disease at a systems level. However, characterising coordinated molecular signatures does not necessarily establish whether the relationships between molecular phenotypes differ across physiological contexts. In particular, how age, sex, BMI and HbA1c jointly modify cross-omic associations, and whether these changes reveal context-dependent molecular organisation, remain insufficiently characterised. Furthermore, identifying such associations alone does not establish the direction of molecular relationships or distinguish candidate directional dependencies from undirected correlations. Integrating context-dependent association analyses with approaches that infer directionally structured molecular relationships may therefore provide a more comprehensive understanding of how physiological state shapes molecular organisation.

Here, we hypothesised that age, sex, BMI and HbA1c influence not only the abundance of individual molecular phenotypes but also the relationships between them, resulting in context-dependent reorganisation of molecular networks across omics layers. To test this hypothesis, we leveraged the IMI DIRECT consortium resource, comprising matched genotype, transcriptomic, proteomic and metabolomic data from 3,027 individuals. First, we characterised the independent associations of each trait with molecular phenotypes across omics layers. We then modelled interactions between traits to identify context-dependent molecular effects and investigated whether cross-omic associations differed according to physiological context. Finally, we applied probabilistic causal inference to investigate directionally structured relationships between age-associated molecules and identify candidate molecular pathways linking upstream and downstream phenotypes. Our analyses revealed widespread associations of physiological traits with molecular phenotypes, context-dependent changes in cross-omic relationships, and directionally structured molecular relationships involving pathways related to mTOR signalling and T2D. Together, these findings highlight the importance of considering physiological context when characterising molecular organisation and provide a framework for investigating the systems-level heterogeneity underlying metabolic disease.

## RESULTS

### BMI, HbA1c, age and sex exert widespread molecular effects across omics layers

To understand the baseline independent effect of the heterogeneous disease context on blood and plasma molecules, we examined associations between molecular phenotypes and T2D related variables in the IMI DIRECT dataset which includes individuals recently diagnosed with T2D and at risk of developing T2D (pre-diabetes status). Molecular phenotypes available included whole blood gene expression (n=16,209), plasma derived targeted proteomics (n=373), and targeted and untargeted metabolites (n=349) from up to 3,029 participants. Binary T2D status, as defined by the study selection criteria (pre-disease, disease), identified no gene expression associations and limited protein and metabolite associations (Tables S1.a; S2-S9). This likely reflects the use of individuals with high diabetes risk as controls and the multi-cohort origin of the dataset. We therefore characterised disease context using quantitative traits; HbA1c levels, a basic criterion for assigning T2D status in the cohort, and BMI, associated with the risk of developing T2D^20^ and also correlated with T2D in the cohort (SFig.1c; ρ = 0.26; p = 10^−44^) to enable a finer resolution of the influence of physiological heterogeneity. Furthermore, we investigated molecular changes with age, a known risk factor for the development of diabetes, and sex, a known biological factor influencing disease context. After multiple testing corrections, all four traits showed extensive, independent molecular associations across transcriptomic, proteomic and metabolomic layers. We identified thousands of associations with a greater proportion of overall molecular phenotypes associated with BMI compared to HbA1c, including 51% (8,256) of genes, 97% (363) of proteins and 83% (288) of metabolites (targeted and untargeted) for BMI and 27% (4,429) of genes, 52% (193) of proteins and 27% (94) of metabolites (targeted and untargeted) for HbA1c. With age, we found a larger proportion of significantly associated molecular phenotypes, including 66% (10,643) of genes, 72% (267) of proteins and 83% (291) of metabolites (targeted and untargeted). Likewise, we identified similar wide-spread effects of sex with the expression of 10,163 genes (63%), 267 circulating plasma proteins (72%), and 299 metabolites (86%)(FDR ≤ 0.05; Tables S1.a; S10-S13, SFig.2a-b). In summary, we found a large number of blood molecular phenotypes independently associated with changes across clinically relevant measurements. Moreover, when controlling for all the other traits, age and sex, as risk factors contributing to disease context and heterogeneity, were associated with more molecular phenotypes than the disease defining traits BMI and HbA1c.

To better understand the biological processes associated with heterogeneous disease context, we performed functional and pathway enrichment analyses across significantly associated molecules for each trait (Tables S14-S25, SFig.3-14). Overall, immune-related processes and changes in the mammalian Target of Rapamycin (mTOR) pathway were enriched among the known functions of blood molecular phenotypes independently associated with disease defining traits, BMI and HBA1c. In particular, for gene expression associated with BMI, we identified regulation of translation and inflammatory response or cell surface receptor signalling pathway, while response to stimulus and other immune-related processes were highly enriched. Pathway enrichment analysis for significantly associated metabolites found the biosynthesis of unsaturated fatty acids and the PI3K-AKT, mTOR and Wnt signalling pathways enriched. For HbA1c-related gene expression and protein levels, we also observed enrichment of immune response (regulation of inflammatory response or neutrophil migration), and of protein processing or protein destabilization. The mTOR pathway was again enriched for metabolites associated with HbA1c, as well as the Advanced Glycation End product and Receptor (AGE-RAGE) signalling pathway. Sex and age associated molecules shared enrichment of multiple immune-related processes; inflammatory responses, leukocyte migration, the regulation of immune effector processes and the MAPK cascade were independently enriched among significantly associated molecular phenotypes with both variables. For metabolites, within the commonly enriched biological pathways for sex and age we found mTOR, PI3K-Akt and Wnt signalling pathways. Sex-related gene expression was also enriched with functions related to monocarboxylic acid metabolic process and response to oxidative stress. In conclusion, we identified the mTOR pathway and multiple immune related processes to be commonly enriched among different molecules independently associated with most traits.

To evaluate our baseline associations and the relevance of our blood-based findings for metabolic diseases, we performed a replication analysis using the gene expression multi-tissue GTEx resource with up to 838 samples across 56 tissues. A previous study evaluated the contribution of BMI, age, sex and diabetes to transcriptomic variation across tissues, identifying a systemic effect^45^. Although our study only includes molecular data from one tissue, we wanted to examine if the larger sample size would identify tissue shared associations with some of the traits. As expected, we found more genes significantly associated with all phenotypes than in any GTEx tissue^45^ (<300). Therefore, we used p-value enrichment analysis (π1)^46^ to compare the distribution of p-values of our significantly associated genes in GTEx, which accounts for sample size differences and aims to estimate the proportion of genes discovered in the IMI DIRECT dataset which were also affected in the GTEx cohort. We found a moderate percentage of genes whose expression was significantly associated with the traits in whole blood, with replications of 33.1% for BMI, 35% for sex, and 34.3% for age, suggesting differences in samples and data processing had strong influence in blood expression analyses. Across all tissues and phenotypes, we found that non-sun exposed skin gene expression showed a higher proportion of shared associations with sex-associated genes in blood than any other tissue-phenotype pair (Table S26; SFig.15). As expected, we found adipose subcutaneous (58%) as the tissue with the larger number of shared associated genes with BMI (Table S26; SFig.15). HbA1c was not recorded in GTEx therefore we compared results with T2D status, given HbA1c was a basic criterion for assigning disease status in the IMI DIRECT cohort and highly correlated with disease status (SFig.1; ρ = 0.66; p = 4.7×10^−231^). Compared to GTEx T2D-associated genes, we found between 0% and 47% shared associations, with nerve tibial tissue sharing the larger number of genes, one of the tissues with a large number of associations in the original GTEx study. Comparison with GTEx differentially expressed genes with sex and age found between 67% to 0% shared associations, with artery aorta showing the largest shared effects with age (59.6%). Overall, we observed a significant linear association between tissue sample size and replication rate for sex (p = 7.1×10^−06^), BMI (p = 5.8×10^−04^), and age (p = 0.015) but not for T2D associated genes with HbA1c genes (p = 0.126). This positive correlation was previously observed for genetic associations^47,48^, with tissues of smaller sample size showing lower replication rates. Overall, our results agreed with previous findings reporting limited shared transcriptional associations to disease traits across tissues.

### Age and sex modulated metabolic disease context response on molecular phenotypes

We identified a moderate number of molecules to be independently associated with more than one disease context variable (BMI and HbA1c) and age, despite both disease traits known to increase with age. The largest proportion of shared molecules was detected for age-BMI associations with proteins (69.7%) and metabolites (69.7%) (SFig.2c-e). The number of molecules significantly associated with both BMI and HbA1c was up to 50.9% for proteins. A three-way comparison identified proteins as the molecules with the greatest percentage of molecules independently associated with BMI, HbA1c and age (37% proteins vs 11.3% expression and 22% metabolites). The higher percentage of proteins associated with T2D related traits was not unexpected, as the proteins assayed in this study were targeted and selected due to their relevance to T2D^49^. Moreover, we expected an additive concordant effect on molecular phenotypes with shared significant associations between BMI-age and HbA1c-age, given that both disease variables are known to increase with age^50,51^. However, we found the direction of effect to be opposite (discordant) between age and BMI for 23% of shared genes, with similar numbers for age and HbA1c (17%) and BMI and HbA1c (12%) (SFig.16). A similar trend was observed for targeted proteins associated with age, BMI and HbA1c (SFig.17). Shared metabolites showed a larger percentage of discordant effects between BMI and age (68%) and HbA1c and age (78%), while only 10% between BMI and HbA1c (SFig.18). In summary, molecules associated with BMI, HbA1c and age often exhibited discordant effect directions, suggesting that the effect of one trait may depend on the state of another.

To identify age-dependent effects of BMI and HbA1c, independently of T2D status, we performed linear regression including an age-by-BMI or age-by-HbA1c interaction term for each molecular phenotype dataset. Significant age-by-BMI interactions identify molecular phenotypes that change with age in a different manner depending on an individual’s BMI. Although age-dependent BMI and HbA1c effects were relatively rare, several metabolites displayed significant context dependence after multiple testing correction (FDR ≤ 0.05; Tables S1.b; S27-S30). We identified a BMI-dependent effect of age for 2 species of sphingomyelin including C26:1 sphingomyelin that was found to increase with age at a greater rate in individuals with a normal BMI compared to individuals with an overweight BMI (p_Age*BMI_ = 4.3×10-4; Fig.1a). Similarly, we identified HbA1c-dependent effects of age for 6 metabolites (FDR ≤ 0.05; Tables S1.b; S31-S34). For example, uridine levels were found to decrease with age only in individuals with lower HbA1c (p_Age*HbA1c_ = 1.3×10-4; Fig.1b).

Given the influence of sex on diabetes development and progression, we next explored the possible sex-specific effects on molecular responses to disease context. Similarly to age, we used interaction terms in linear regression models for sex-by-BMI or sex-by-HbA1c interactions. We found significant sex-specific effects of BMI for the expression of 2 genes, 83 proteins and 94 metabolites (FDR ≤ 0.05; Tables S35-S38, S80.b). Our results included examples such as the expression of *TPPP3* (tubulin polymerization promoting protein family member 3) that was found to significantly increase with higher BMI only in men (p_Sex*BMI_ = 5.9×10^−7^; Fig.1c). In a similar manner, levels of creatine increased with higher BMI only in men (p_Sex*BMI_ = 1.2×10^−6^; Fig.1e). Moreover, levels of leptin (LEP) significantly increased with higher BMI in both men and women, yet with a higher rate for men (p_Sex*BMI_ = 7×10^−13^; Fig.1d). We also observed sex-specific effects of HBA1c for 6 proteins and 4 metabolites (FDR ≤ 0.05; Tables S39-S42). For example, levels of protein STK4 (serine/threonine kinase 4) significantly decreased with higher HbA1c only in women (p_Sex*HbA1c_ = 2×10^−4^; Fig.1f), while levels of the metabolite proline increased with HbA1c, also only in women (p_Sex*HbA1c_ = 1.4×10^−3^; Fig.1g). Overall, we observed a larger sex-dependent effect on the molecular response to metabolic disease context than with age, despite their similar number of independent associations reported for each of the molecular phenotypes.

### BMI modified HbA1c-associated protein signatures linked to T2D risk

Defining disease context by quantitative values allowed us to identify molecular responses to glycaemic dysregulation that depend on BMI, uncovering putative causal proteins for diabetes subtypes. We further investigated the joint influence of BMI and HbA1c on molecular phenotypes, and tested for HbA1c-by-BMI interactions, identifying 8 proteins whose levels were associated with HbA1c levels but behaved differently depending on the individual’s BMI (Tables S1.b; S43-S46). These were LEP, FABP4, IGFBP2, IGFBP1, PON3, FETUB, SRCRB4D, and ANG. Levels of LEP (leptin) were positively associated with HbA1c in individuals with low BMI, whereas individuals with high BMI showed leptin levels negatively associated with HbA1c (Fig.1h). In another example we detected that IGFBP2 (insulin-like growth factor binding protein 2) and HbA1c were negatively associated, with the effect size of this association (β_HbA1c_) decreasing with BMI. As a result, individuals classified as obese had seemingly similar levels of IGFBP2 across levels of HbA1c, while those classified with normal BMI showed an expected decrease in IGFBP2 as HbA1c increased^52^ (Fig.1i). Out of these 8 proteins, 6 were also associated with BMI, age and HbA1c: leptin, FABP4 (fatty acid binding protein 4) and PON3 (paraoxonase 3) were independently associated with different directions of effect while IGFBP2, FETUB (fetuin B) and SRCRB4D (scavenger receptor cysteine rich family member with 4 Domains) showed the same direction of effects across variables (SFig.17). For the other two, ANG (angiogenin) was only significantly associated with BMI (p_BMI_ = 7.2×10-4) and IGFBP1 with both BMI and age (p_BMI_ = 2×10-121; p_age_ = 1.2×10-9). Furthermore, 7 out of these 8 proteins have been previously associated with Polygenic Risk Scores (PGS) for T2D^53^, highlighting their potential role in mediating T2D genetic risk. For all of them, the authors also found that BMI mediated the effect of the PGS on circulating levels of the proteins, supporting a context dependent effect in which the level of the proteins was dependent on both BMI and a disease status. On the whole, we showed that obesity can change the molecular response to glycaemic dysregulation based on eight proteins, seven of which had been previously reported to play a role in BMI-mediated T2D risk and with SRCRB4D as a novel finding.

**Figure 1:**
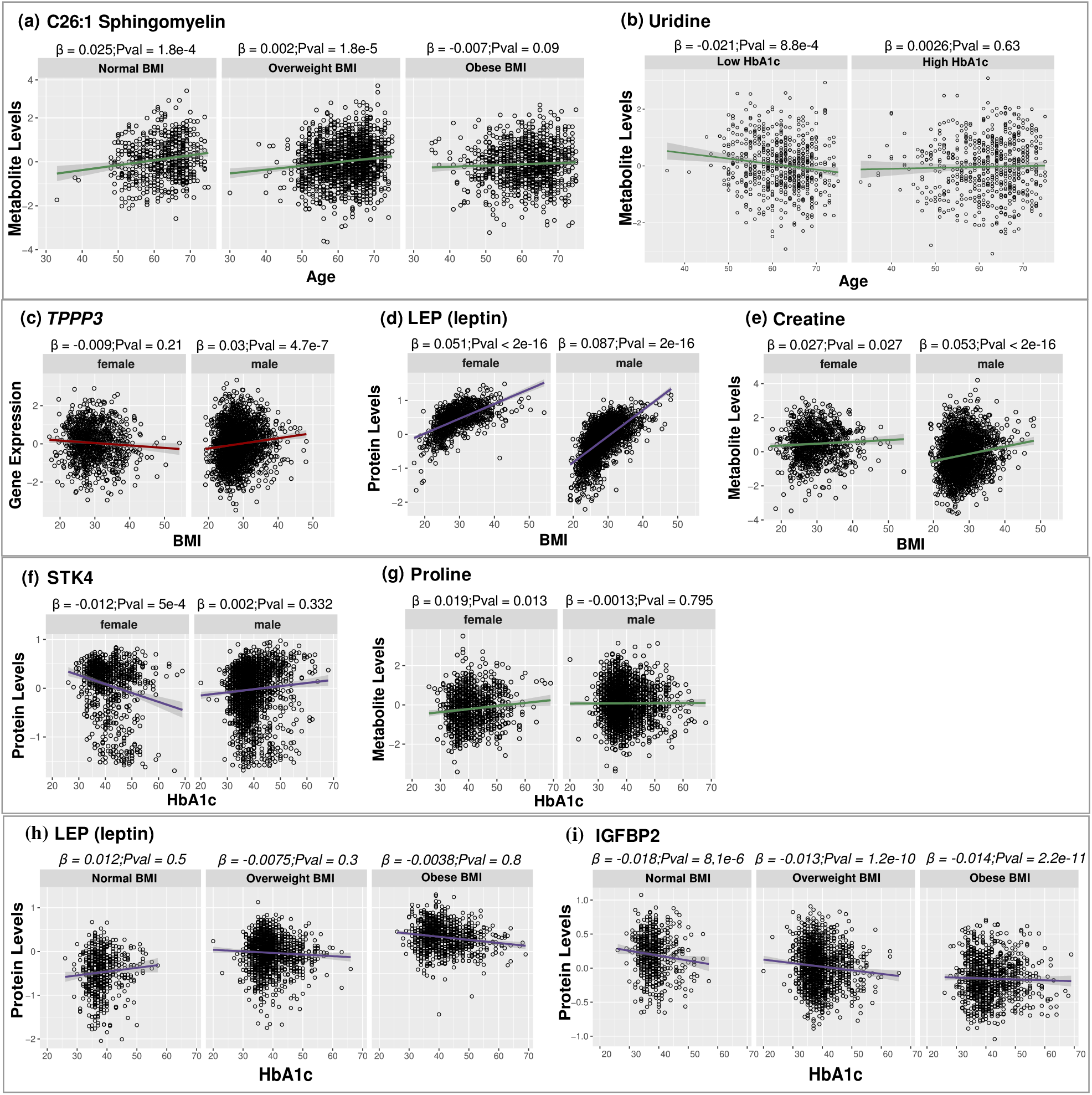
Significant context dependent effects on molecular phenotypes. Age-dependent metabolic disease context response: (a) Metabolite C26:1 sphingomyelin (SM.C24.1) was found significant for an age-dependent effect of BMI. C26:1 Sphingomyelin levels against age plotted separately for individuals in different BMI classification groups. (b) Metabolite uridine was found significant for an age-dependent effect of HbA1c. Uridine levels plotted against age for the 20% of individuals with the lowest HbA1c and the 20% of individuals with the highest HbA1c in the cohort; The range of HbA1c levels in the cohort was 20-69 mmol/mol with a mean of 39.7 mmol/mo. Sex-specific metabolic disease context response: (c) Gene *TPPP3* was found significant for a sex-specific effect of BMI; gene expression of *TPPP3* against BMI plotted separately for female and male individuals. (d) Protein LEP (leptin) was found significant for a sex-specific effect of BMI; Levels of leptin against BMI plotted separately for female and male individuals. (e) Metabolite creatine was found significant for a sex-specific effect of BMI; Levels of creatine against BMI plotted separately for female and male individuals. (f) Protein STK4 was found significant for a sex-specific effect of HbA1c; Levels of STK4 against HbA1c plotted separately for individuals with low and high levels of HbA1c. (g) Metabolite proline was found significant for a sex-specific effect of HbA1c; Levels of proline against HbA1c plotted separately for female and male individuals. BMI-dependent effects of HbA1c: (h) Protein LEP (leptin) was found significant for BMI-dependent effect of HbA1c. Levels of leptin against HbA1c plotted separately for individuals in different BMI classification groups. (i) Protein IGFBP2 was also found significant for BMI-dependent effect of HbA1c; Levels of IGFBP2 against HbA1c plotted separately for individuals in different BMI classification groups.

### Molecular responses to ageing diverged between men and women

Because ageing and sex jointly influence metabolic disease risk, we asked whether molecular responses to ageing differ between men and women. Firstly, we identified a 75% overlap between age and sex associations shared across molecules, suggesting widespread sex-specific effects of age (SFig.2c-3; 19-21) Secondly, we examined sex-specific effects (sex-by-age interactions) to detect molecular phenotypes that change with age in a different manner in men compared to women. After multiple testing corrections, significant sex-specific effects of age were observed for 850 genes, 84 proteins and 64 metabolites (FDR ≤ 0.05; Tables S1.b, S47-S50). For example, the expression of *CDKN1A* (cyclin dependent kinase inhibitor 1A) showed a sex-specific effect of age with an increase in the gene’s expression with age only for women (p_Age*Sex_ = 2.7×10^−2^; Table S47; Fig.2a). In addition, we observed certain cases where sex-specific effects of age showed opposite direction of effects that were not detectable as associations with age and/or sex (SFig.22-24). For instance, gene expression of *NARF* (nuclear prelamin A recognition factor) was neither associated with age nor sex (Table S10), yet showed a sex-specific change with age with opposite direction of effect, detectable when considering an interaction term (p_Age*Sex_ = 7.2×10^−3^; Table S47; Fig.2b). Proteins found significant for a sex-specific effect of age included SOST, GDF15 and IL6 (p_Age*Sex_< 0.02; Table S48). For example, we observed that the glycoprotein SOST (sclerostin) increased with age only for male individuals (p_Age*Sex_ = 2×10^−4^; Fig.2c), an effect previously observed^54^. Moreover, IL6 (Interleukin-6) showed a significant increase in its levels with age only in men (p_Age*Sex_ = 2×10^−4^; Fig.2d). Levels of metabolites choline, uridine, piperine, propionylcarnitine (C3) and creatine also showed sex-specific effects of age (p_Age*Sex_<0.006; Tables S49-S50). For instance, choline levels, previously linked with older age and unfavourable cardio-metabolic risk profiles^55^, increased with age only in men (p_Age*Sex_ = 0.01; Fig.2e), while levels of creatine were associated with sex but not with with age (Table S13) and were also found to decrease with age only in men (p_Age*Sex_ =1.2×10^−3^; Fig.2f). These results identify sex-specific ageing as the most pervasive context-dependent effect observed in this study, suggesting that molecular ageing trajectories diverge substantially between men and women.

**Figure 2:**
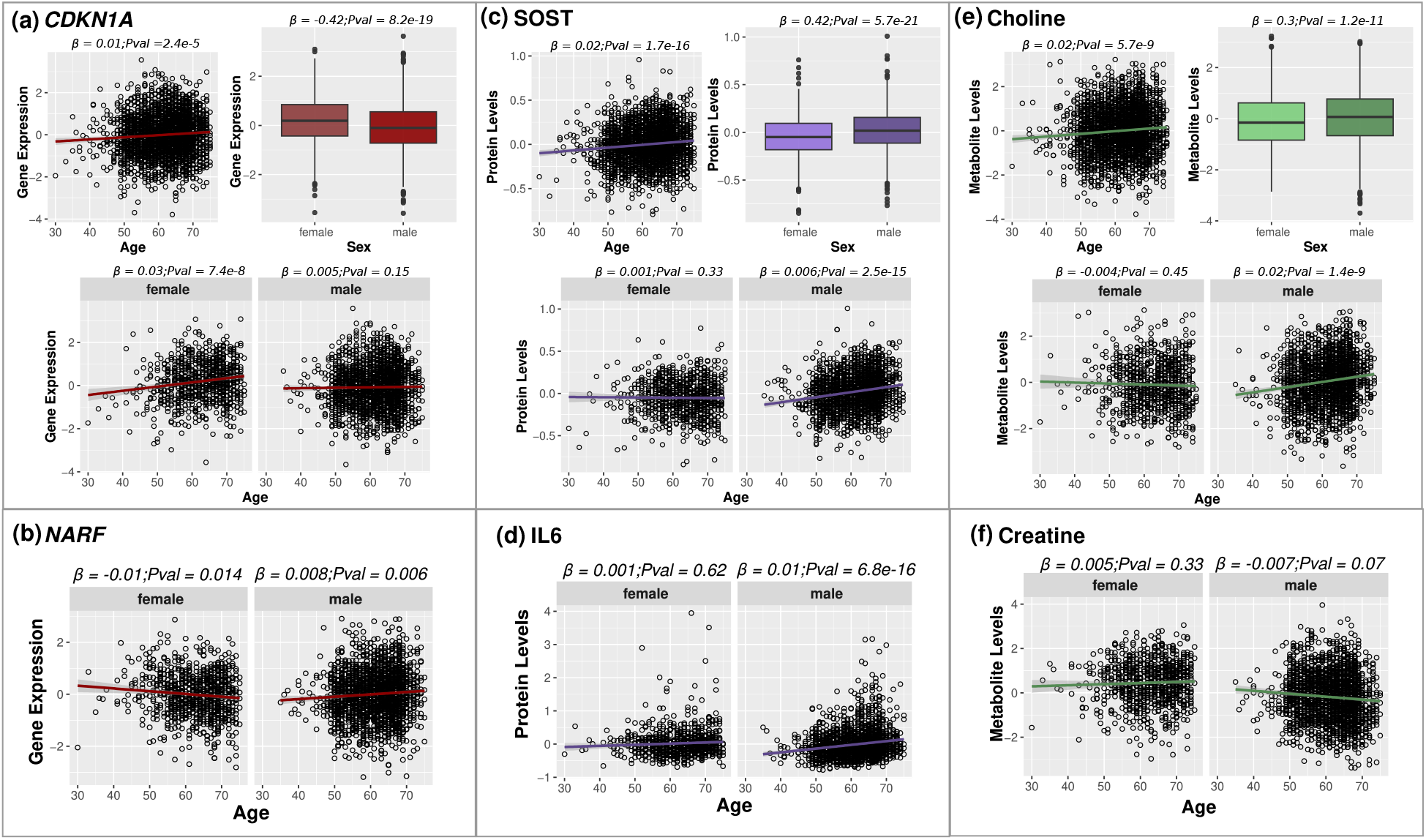
Significant sex-specific effects of age on molecular phenotypes. (a) Expression of the gene *CDKN1A* (coding for the p21 protein) was independently associated with age and sex and showed a sex-specific effect of age. Gene expression of *CDKN1A* against age plotted separately for female and male individuals. (b) Gene *NARF* was found significant for a sex-specific effect of age. Gene expression of *NARF* against age plotted separately for female and male individuals. (c) SOST protein levels were independently associated with age and sex and showed a sex-specific effect of age. Levels of protein SOST against age plotted separately for female and male individuals. (d) Protein IL6 (interleukin-6) was found significant for a sex-specific effect of age. Interleukin-6 levels plotted against age separately for female and male individuals. (e) Levels of metabolite choline were independently associated with age and sex and showed a sex-specific effect of age. Levels of choline plotted against age separately for female and male individuals. (f) Metabolite creatine was found significant for a sex-specific effect of age. Creatine levels plotted against age separately for female and male individuals.

### Metabolic disease traits rewired cross-omic molecular relationships

Our results have shown a widespread heterogeneous effect of biological and clinical variables on multiple molecular phenotypes, many of which replicate previous findings in other tissues, and suggest intrinsic and extrinsic exposures may alter the relationships between molecules. Therefore, we investigated whether physiological and metabolic traits influence not only molecular abundance but also the relationships between molecules. Firstly, we used linear models for pairs of different types of molecules (gene expression, proteins and metabolites) to identify associations where levels of one molecule depend on the levels of another, independently of these disease traits. We found 1,573,779 significant associations (13% of all tested pairs) between 16,927 molecules (99.9% of tested molecules; FDR ≤ 0.05; Tables S1.c, S51-S55; Fig.3a). The number of pairs found to be significantly associated were the lowest between gene expression and metabolite levels (8%), followed by 15% of protein-metabolite pairs. For gene expression-protein pairs, 18% out of the 6,045,957 possible pairs were significantly associated, reflecting a closer relationship between the processes regulating transcription and translation compared to metabolites production.

**Figure 3:**
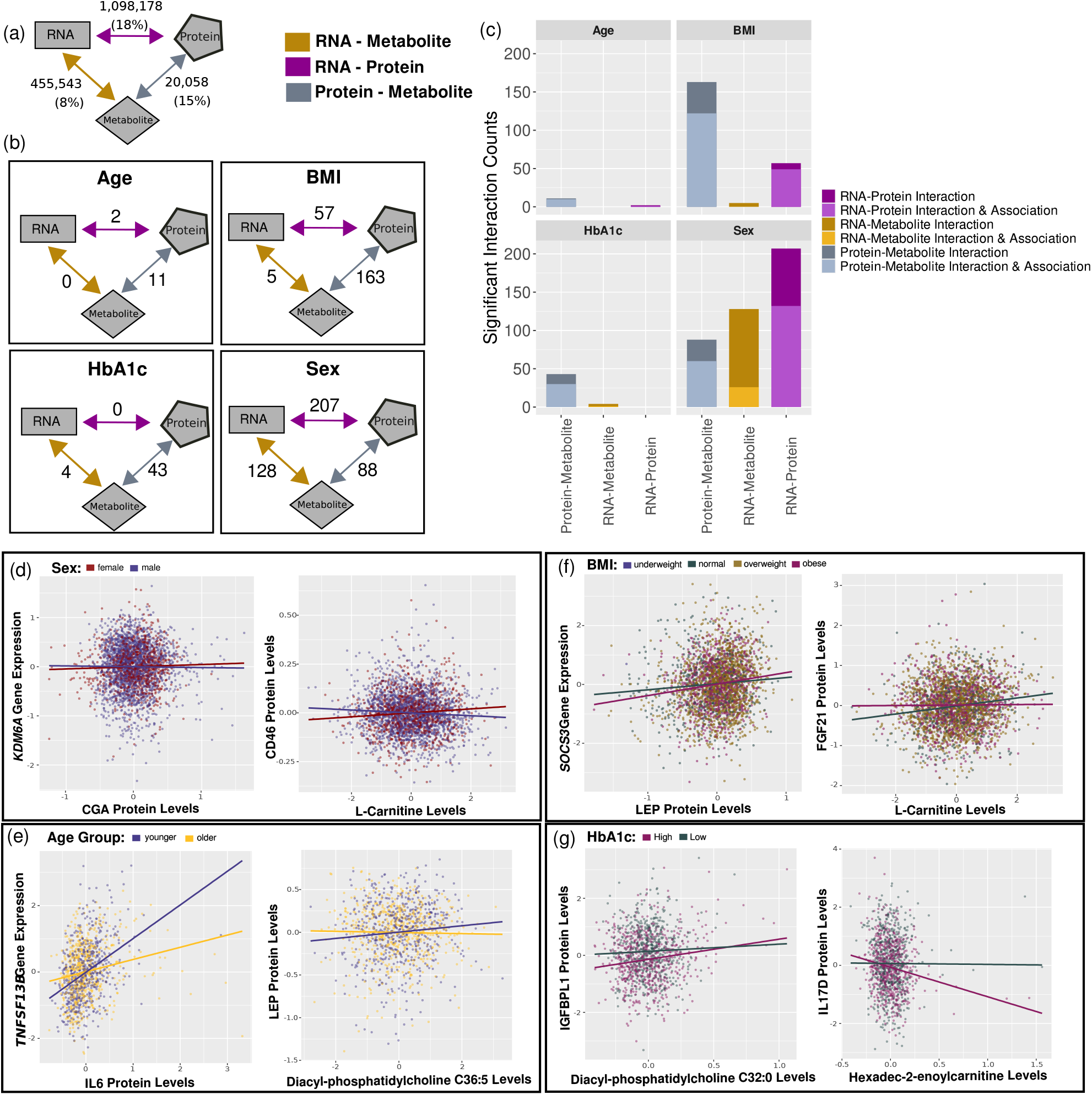
Significant associations (relationships) across molecules and context dependent effects (interactions) on molecule associations: (a) Counts of associations across molecules; a total of 1,573,779 associations were identified (13% of all tested pairs, FDR ≤ 0.05) between 16,927 molecules (99.9% of tested molecules). (b) Counts of interactions across molecules for age, BMI, HbA1c levels and sex respectively. (c) Counts of molecular phenotype relationships found significant for a context dependent effect; on each bar, counts of pairs found significant for both an association between the two molecules and a context dependent effect modulating this relationships is depicted; association and interaction were assessed from the same model. Examples of context dependent effects (interactions) modulating molecule associations: (d) Sex interactions: the relationship between KDM6A gene expression and CGA protein levels and the relationship between CD46 protein levels and metabolite L-Carnitine levels behaved differently between male and female individuals. (e) Age interactions: the relationship between TNFSF13B gene expression and IL6 protein levels and the relationship between LEP protein levels and metabolite Diacyl-phosphatidylcholine C36:5 levels behaved differently in younger and older individuals; the 20% of individuals with the lowest age (younger) and the 20% of individuals with the highest age (older) in the cohort were plotted. (f) BMI interactions: the relationship between SOCS3 gene expression and LEP protein levels and the relationship between FGF21 protein levels and metabolite L-Carnitine levels behaved differently depending on individuals’ BMI; for visualisation purposes linear regression lines are depicted only for individuals with normal and obese BMI. (g) HbA1c interactions: the relationship between IGFBPL1 protein levels and metabolite Diacyl-phosphatidylcholine C32:0 levels and the relationship between IL17D gene expression and metabolite Hexadec-2-enoylcarnitine levels behaved differently in individuals with high and low levels of HbA1c; the 20% of individuals with the lowest HbA1c and the 20% of individuals with the highest HbA1c in the cohort were plotted.

Subsequently, we examined the influence of sex, BMI, HbA1c and age on the pairwise associations between molecules and found a higher proportion of molecular phenotype pairs to have a dependent relationship with metabolic disease relevant traits. Interaction models for every molecule pair were assessed for sex, BMI, HbA1c or age dependence in their relationships (Table S1.d). Sex and BMI had the strongest influence on molecular relationships; sex was more likely to influence gene expression relationships with proteins and metabolites while BMI was more likely to influence relationships between proteins and metabolites (Fig.3b). Moreover, when assessing the full interaction models, we observed additional context dependent effects modulating their relationship that were present without an independent, non-disease related, association between them (84 gene expression-protein, 110 gene expression metabolite and 83 protein metabolite pairs; Fig.3c). For example, the gene expression-protein relationship between *KDM6A* (Lysine Demethylase 6A) and CGA (Glycoprotein Hormones, Alpha Polypeptide) and the protein-metabolite relationship between CD46 (membrane cofactor protein - MCP) and L-Carnitine was different between male and female individuals (Fig.3d). In addition, we observed that BMI influenced the relationship between *SOCS3* (Suppressor of Cytokine Signaling 3) gene expression and LEP protein levels and the relationship between protein FGF21 (Fibroblast Growth Factor 21) and L-Carnitine levels (Fig.3f). Similarly, HbA1c levels influenced the relationship between IGFBPL1 (Insulin Like Growth Factor Binding Protein Like 1) protein levels and Diacyl-phosphatidylcholine C32:0 levels and the relationship between IL17D (Interleukin 17D) protein levels and Hexadec-2-enoylcarnitine levels (Fig.3g). Age influenced a smaller number of pairs compared to the other traits (Tables S71-S75; Fig3b-c). Examples include, the relationship between *TNFSF13B* gene expression and IL6 protein levels and LEP protein levels and Diacyl-phosphatidylcholine C36:5 levels that behaved differently in younger and older individuals (Fig.3e). Overall, protein-metabolite pairs were influenced to a higher proportion by disease relevant traits compared to pairs involving gene expression and other molecules (Table S1.d). In addition, when assessing the joint influence of the different metabolic traits, we observed that sex, BMI, HbA1c and age effects were most often unique/not shared with each other (SFig.25).

Context dependent effects on molecular relationships suggests that the effects of a trait on one molecule propagate to other molecules. To investigate the directionality of molecular relationships, we leveraged age as an exogenous quantitative variable whose value is not influenced by the molecular phenotypes under study. Because the direction of influence between age and molecular phenotypes is biologically established, age was used to inform Bayesian network analysis to determine directionality of effect. Therefore, we tested whether age independently affected pairs of associated age-related molecules or whether its effects on a given molecule were mediated through another molecule. Here, we investigated relationships between pairs of molecules of different types and extended the analyses to those of the same type, and identified 97,807,420 significant associations (84% of all tested pairs) between 11,169 (99.7%) age-related molecules (FDR ≤ 0.05; Tables S1.e, S76; SFig.26a-b). For these associations, we evaluated the three possible models (SFig.26c): models 1 and 2 where age affects one of the molecules and then, in turn, that molecule influences the levels of the other (or vice versa) and a model 3 where age affects both molecules independently. The best fit across the three models was evaluated using the Bayesian Information Criterion (BIC) score for each model, hereby noted as dependent (models 1 and 2) and independent models (model 3). For every age and molecule pair trio, we considered that there was substantial evidence supporting the best fit if one of the three models had a BIC score difference ≥ 6 (strong evidence supporting the model) with either of the other two models.

We were able to get a supported decision for the best fit model for 91% of associated, age-related molecular phenotype pairs (Table S77; SFig.26d-e). Of those, we observed that for pairs of the same molecule type, age was more likely to independently affect the two: 79%, 70% and 98% of age-related pairs for gene-gene expression, protein-protein and metabolite-metabolite levels, respectively. In contrast, the same model where age was more likely to independently affect the two molecules was only observed in 11%, 8% and 20% of the gene expression-protein, gene expression-metabolite and protein-metabolite pairs. Overall, we found that for 66% of gene-protein pairs, age influenced the expression of the gene and that in turn influenced the levels of the protein. For gene-metabolite pairs we found that for 43%, age influenced expression with consequences for metabolite levels, for 48% the relationship was reversed, and for the remaining 9% the age effects were independent. Finally, for 56% of protein-metabolite pairs, we found that age influenced metabolite levels and that in turn influenced the levels of the protein. Altogether, for pairs of molecules that best fitted one of the dependent models we were able to assign a direction in their relationship, often with a similar proportion of trios assigned to each direction.

### Age-associated molecular networks highlight mTOR signalling and T2D-related pathways

To visualise the landscape of age-associated molecular changes, we constructed a network using age as an exogenous variable and incorporating molecular relationships with inferred directionality. The resulting network comprised 11,166 molecules as nodes and ∼1.1×10^6^ edges with directionality inferred based on the causal framework developed in the previous section (S78). The network identified highly connected hubs enriched for immune and metabolic regulators and converged on pathways implicated in ageing and metabolic disease, including mTOR signalling. We observed that 45% of molecules were connected with more than 100 nodes (degree > 100). The most central element of this age-network was the protein interleukin 6 (IL6), connected with 9,015 molecules, followed by proteins LILRA5 (leukocyte immunoglobulin like receptor A5) and PLAUR (plasminogen activator, urokinase receptor) (degree > 8,000). Using this network as a template to explore the effects of metabolic disease relevant traits, we focused on the mTOR signalling pathway and on a sub-network relevant for T2D through genetic evidence.

The mTOR signalling pathway was enriched in our own analyses and has been characterised as a central regulator of metabolism, also implicated in age-associated processes such as loss of proteostasis and cellular senescence with its inhibition suggested to extend life span and delay the onset of age-associated diseases^56–58^. A sub-network was constructed including the 35 nodes (34 genes and 1 protein) from the mTOR pathway that were significantly associated with age and had evidence supporting a direction of effect between molecules, as well as their first neighbour molecules (Fig.4; S78). Many of the 35 mTOR nodes mediated the effect of age towards multiple molecules. However, central elements of the network such as the protein PGF (Placental Growth Factor) were modulated by multiple other age-related molecules, as the direction of effect of age was reported affecting PGF. In our study, the levels of PGF increased with age (β_Age_ = 0.03; p_Age_ = 2.7×10^−26^) while also being higher in men (β_male_ = 0.47; p = 7.3×10^−26^). In addition, levels of the protein have been shown to be higher in post-menopausal compared to pre-menopausal women, suggesting that this sex bias could be mediated by hormones^59^. While age-related changes in the levels of PGF were mostly mediated by other molecules, its levels were also mediating some proteins such as GDF15. In our network, age-related changes in GDF15 levels were mediated by the effect of age on 11 components of the mTOR pathway including protein levels of PGF and gene expression of the genes *PRKAA1*, *RPS6KB1 PIK3R2*, *AKT3*, *MAPK3* and *MTOR* (Fig.4). Other genes highly connected in our network were *PIK3R2* and *AKT3,* both part of the PI3K-AKT signalling pathway which has known implications in T2D^60^. From our age-by-sex interaction results, we observed that out of the 35 molecular components of mTOR in the network, gene expressions of *MAPK1*, *PIK3CD* and *ULK1* were significant for a sex-specific effect of age (Table S47). In addition, levels of 22 proteins in the network, including GDF15 and IL6 and levels of metabolite uridine were also significant for a sex-specific effect of age (Tables S48, S50). Moreover, protein TNFRSF1B was found to have a sex-dependent relationship with 89 genes including *SORT1*, *TCF19*, *TNFSF13B*, *CD274* and *CASP1* (Table S56) and a BMI-dependent relationship with 20 genes including *SORT1*, *CFB* and *LGALS9* (Table S61). Finally, proteins IGFBP1, IGFBP2, LDLR and GPR64 showed a BMI-dependent relationship with multiple choline species (Table S64). Overall, these findings highlight the mTOR pathway as a central component of the age-associated molecular network, illustrating how age-related changes in molecular phenotype levels are interconnected and how these relationships, further shaped by sex and BMI, may contribute to the remodelling of metabolic regulation across the lifespan.

**Figure 4:**
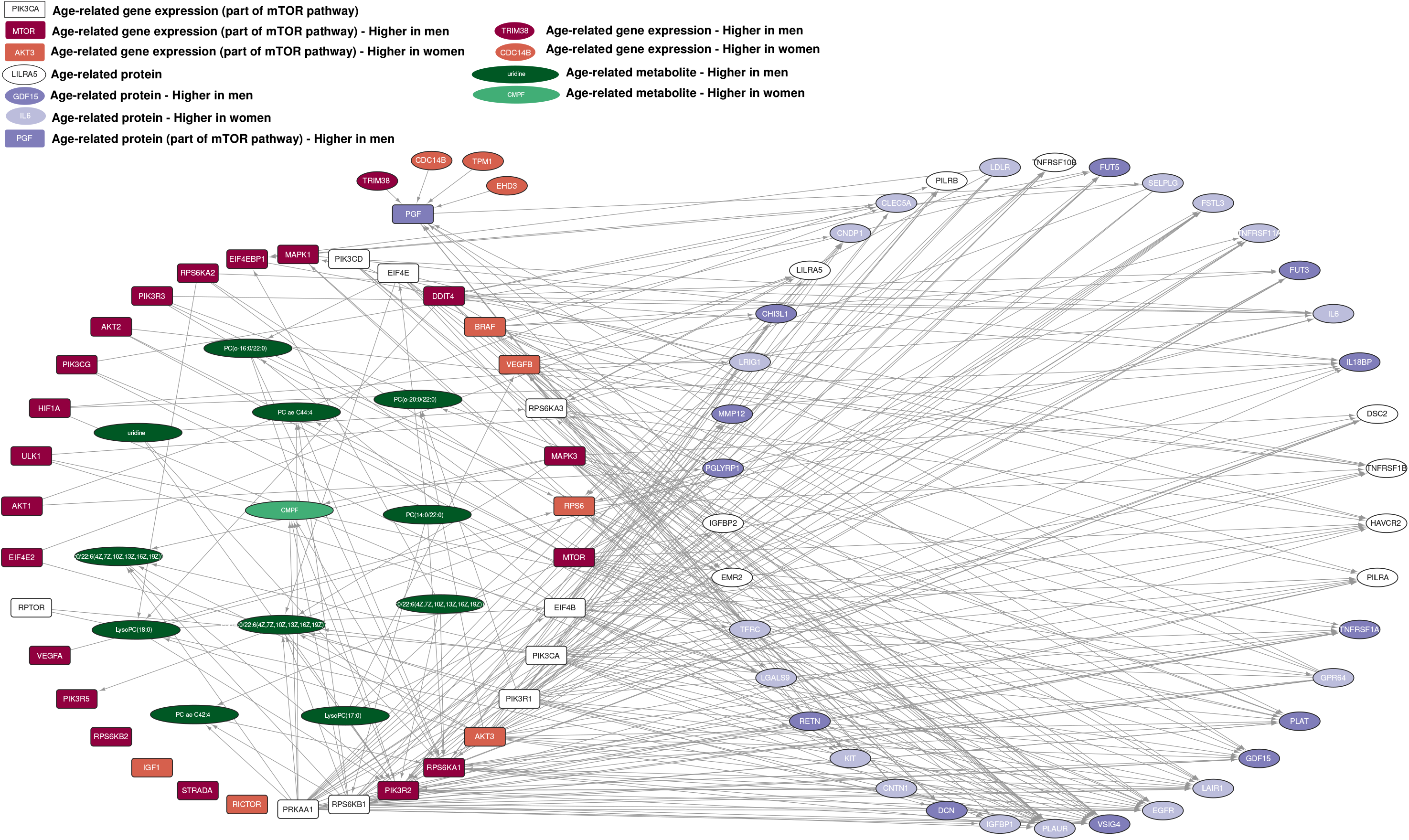
Network of age-related molecular phenotypes that are part of the mTOR signalling pathway (35 molecules) and their 1st neighbours. Square nodes signify molecules that are part of the mTOR signalling pathway. Only edges indicating a significant association between molecules with FDR ≤ 10^−11^ were included. Coloured nodes highlight molecules that were also associated with sex: gene expression in red, proteins in purple and metabolites in green. Darker nodes indicate that the levels of the molecule were higher in men while lighter nodes indicate that levels were lower in men compared to women. Direction of edges was assigned using the causal inference results (BIC score ≥ 6) indicating associations between molecules were the effect of age on the target node was mediated by the source node or vice versa.

To explore molecular relationships associated with T2D, we identified relevant molecules using a colocalisation analysis between genetic loci associated with age-related molecules (Quantitative Trait Loci; QTLs identified by Brown et al.^47^) and loci associated with T2D from a GWAS study^16^. We identified 7 age-related genes whose expression Quantitative Trait Loci (eQTLs) colocalised with genetic variants associated with T2D in our dataset (Tables S79-S86). To visualise the relationships between these molecules and other age-related molecules we focused on a subset of our network around them (S78). In addition, we included the SNPs that were found to influence the levels of the 7 genes identified by the colocalisation analyses resulting in a network of 67 nodes and 1,211 edges (Fig.5a). Among the genes colocalised with T2D loci we found *PRUNE* (prune exopolyphosphatase 1), *POGZ* (pogo transposable element derived with ZNF domain), *RFX5* (regulatory factor X, 5 influencing HLA Class II expression) and *RAB1A* (member RAS oncogene family) which had 80 connections with other proteins and metabolites. Metabolites in the network included a phosphatidylcholine, a lysophosphatidylcholine and three species of carnitine, an expected observation since low levels of L-carnitine have previously been associated with various diabetic complications^61^. Proteins influenced by the expression of genes associated with T2D included LEP, PON3, IGFBP1 and IGFBP2, which were also found to be significant for an HbA1c-by-BMI interaction. Levels of IGFBP2 in particular, were found to be influenced by 3 out of 7 genes with a colocalised variant for T2D. Finally, many relationships between molecules in the network were found to be modulated by BMI, HbA1c, age and sex (Fig.5b, Fig.3f-g). For instance, the relationships between L-Cartine and proteins FGF21 and DPP4 as well as the relationships between *SORT1* and TNFRSF1B, and *SOCS3* and LEP were influenced by BMI. In a similar manner, HbA1c was found to influence the relationships between ILD17D and trans-Hexadec-2-enoyl carnitine, and DPP4 and L-Histidine. Our networks demonstrate how metabolic disease traits have the potential to modify, strengthen or weaken the baseline effects of one molecule over another, potentially altering the downstream consequences that define disease response.

**Figure 5:**
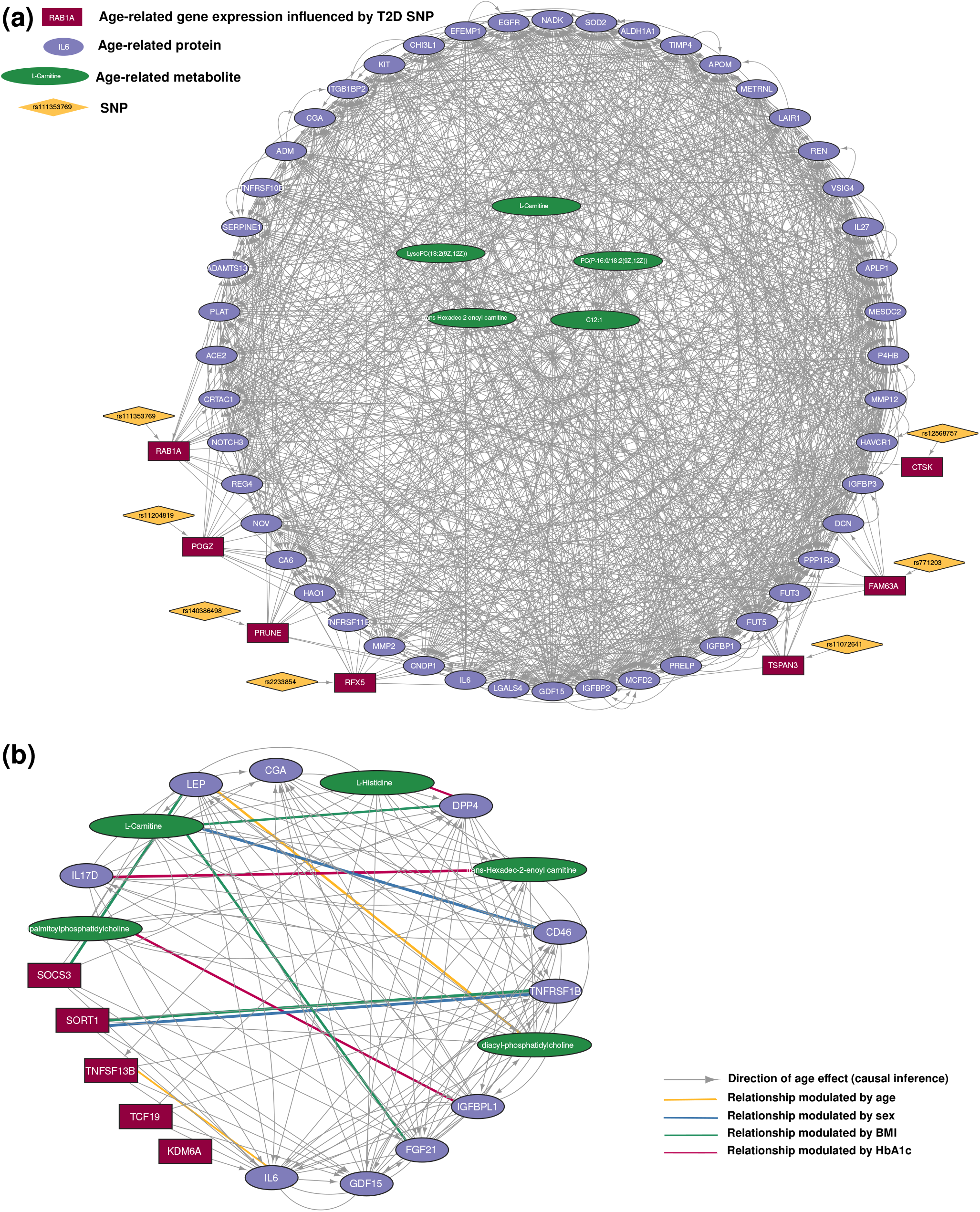
(a) Network of age-related molecular phenotypes and genetic associations that are implicated in Type 2 Diabetes (T2D) based on colocalisation results with GWAS loci associated with T2D. Shape is informative of colocalisation results: diamond nodes show SNPs that influence the levels of colocalised gene expressions and square nodes show genes whose expression was influenced by SNPs colocalised in both GWAS studies (7 genes; unadjusted and adjusted for BMI). Circular nodes show molecules (proteins and metabolites) associated with genes influenced by SNPs colocalised with GWAS loci associated with T2D (1st neighbours). Colour indicates node type: red for gene expression, purple for proteins, green for metabolites and yellow for SNPs. Direction of edges was assigned using the causal inference results (BIC score ≥ 6) indicating associations between molecules were the effect of age on the target node was mediated by the source node and vice versa. Edges with no direction represent associations between molecules were the effect of age best fitted the independent model. For genetic effects, direction was assigned from SNP to molecule as the only possible direction of effect. (b) Sub-network where the impact of BMI, HbA1c, sex and age on the relationships of molecules is shown based on some of the context-dependent effects identified. Context-dependent effects were examined across different types of molecules; effects depicted here are the ones also shown in figure panel 3d-g. The levels of these molecules are associated, yet several relationships were found to be different in younger compared to older individuals (yellow line), between men and women (blue line), between individuals with different BMI (green line) or individuals with different HbA1c levels (pink line).

## DISCUSSION

In this work we aimed to characterise the joint molecular effects of sex, age, BMI and HbA1c, traits used in the diagnosis and stratification of T2D, across multiple molecular layers. We identified widespread effects of these traits across transcriptomic, proteomic, and metabolomic phenotypes, while also uncovering hundreds of context-dependent effects that were not captured by additive models alone. Recent studies examining the contribution of individual traits such as sex, age, and ethnicity to gene expression variation have largely reported additive and tissue-specific effects^45^. In contrast, our findings demonstrate that the molecular response to a heterogeneous metabolic environment is highly context dependent, with the molecular effect of one trait frequently modified by the state of another. These observations indicate that a substantial component of biological variation may remain obscured when physiological traits are analysed independently, supporting a view of metabolic disease as a collection of heterogeneous and interacting physiological states in a continuum rather than discrete static categories.

Examples of these non-additive effects included the proteins leptin and IGFBP2, both of which have established links to obesity and T2D. Previous studies have reported BMI-dependent relationships between leptin and HbA1c in individuals with T2D^62^, while IGFBP2 has been shown to be inversely associated with BMI, regulated by leptin levels, and implicated in obesity prevention and diabetes treatment^63,64^. Our analyses extend these observations by demonstrating that such known relationships can themselves be jointly modified by metabolic disease relevant traits. In particular, IGFBP2 exhibited distinct responses to age depending on sex (SFig.23), and to BMI depending on HbA1c (Fig.1i), suggesting that its role in metabolic disease may differ depending on obesogenic and glycaemic context. Moreover, the regulation of IGFBP2 by leptin levels may be challenged, as causal inference analyses supported a model in which IGFBP2 mediated the effect of age on leptin levels and not the other way around (Fig.5b). A recent study has identified both proteins as potential biomarkers of T2D based on multiple polygenic risk scores (PGS), including PGS partitioned by obesity related traits^53^. Together, these findings suggest that the value of molecular biomarkers and therapeutic targets may depend strongly on physiological state, highlighting the importance of incorporating age, sex, adiposity, and glycaemic status into biomarker interpretation and patient stratification.

The disease-related variables used to define context in our analyses are themselves widely used to diagnose or characterise T2D subtypes, with direct implications for treatment selection and disease management. Multiple subtype frameworks based on clinical variables^15^, genetic architectures^18,65^, and integrated molecular phenotypes^14^ have consistently demonstrated substantial heterogeneity within T2D. However, evidence supporting the clinical utility and long-term stability of these subtype classifications remains limited^66^. This limitation likely reflects both differences in genetic predisposition and environmental exposure, as well as the inherently dynamic nature of metabolic disease progression over an individual’s lifetime^67^. Our results show that there are potentially very different, if not unique, molecular profiles associated with the disease-context variables. Disease subtype classification and therapeutic response may depend on underlying molecular states that shift as glycaemic control, adiposity, and other physiological variables change over time. This supports previous proposals advocating dynamic biomarker assessment and periodic disease reclassification in metabolic disease^67^.

Beyond the responses of individual molecular phenotypes, our analyses demonstrate that cross-omic molecular relationships are themselves dynamic and condition dependent. We identified extensive context-dependent effects on the relationships between different types of molecules (transcripts, proteins, and metabolites), consistent with known biological processes including transcript-protein discordance, post-transcriptional regulation, and metabolic buffering. These findings indicate that metabolic relevant traits reshape not only molecular abundance, but also the coordination and directional organisation of molecular systems across omics layers. By integrating probabilistic causal inference with cross-omic association networks, we further identified directionally structured molecular relationships influenced by age and physiological state. Notably, dependent models dominated the effects of age between molecules of different type, showing that the effect of age was more likely to be mediated from one omics layer to another (SFig.26), consistent with our previous work demonstrating how genetic effects propagate across molecular phenotypes in a hierarchical manner^47^. Together, these observations support a model where metabolic disease response emerges through coordinated rewiring of interconnected molecular systems rather than isolated perturbations of individual molecules.

## METHODS

### Cohort characteristics

We employed a subset of the previously published IMI DIRECT cohort dataset. Individual recruitment information, molecular phenotype generation and data collection is fully described in Koivula et al.^68,69^ and Brown et al.^47^. Here, we offer a summary for clarity:

The data is derived from two parallel cross-sectional cohorts of European ancestry adults, recruited from 7 centres across Europe. The first cohort (Working Package 2.1; WP2.1) includes individuals with blood glucose concentrations within the normal glucose control and fasting Haemoglobin A1C (HbA1c) relevant for pre-diabetes status, according to the American Diabetes Association criteria^70^, while the second cohort (Working Package 2.2; WP2.2) includes individuals recently (within 6–36 months of study enrolment) diagnosed with T2D according to ADA criteria^71^. The age range of individuals was 37-85 years, the ratio of male to female individuals was approximately 3:1 and their BMI range was 17-54. The average BMI of individuals was an overweight mean BMI of 28.7. T2D status overlapped with the cohort membership (WP2.1 and WP2.2) for all but 105 of the participants, therefore any technical differences due to sample processing between cohorts were also highly correlated with T2D status (ρ = 0.92; p < 2.2×10^−308^) and HbA1c levels (ρ = 0.66; p = 1.2×10^−225^; SFig.1). Of the 3,029 individuals included in DIRECT, only those with complete data for age, sex, BMI and HbA1c were included in this study, resulting in N= 3,027 samples (2,234 assigned a pre-diabetic and 793 assigned a diabetic status).

### Data collection and pre-processing

Genotyping for the all samples was performed using the Illumina HumanCore array (HCE24 v1.0), called using the GenCall algorithm. Sample and ethnic outliers filtering, genotyping quality control (QC) and Imputation to the 1000 Genomes Phase 3 CEU reference panel were performed by Brown et al.^47^.

RNA sequencing was performed using the Illumina HiSeq 2000 platform and 49 bp paired-end reads. Reads were mapped to the GRCh37 reference genome^72^, and gene quantifications were calculated as Fragments Per Kilobase of transcript per Million mapped reads (FPKM). Genes with more than 50% of zero reads, genes from chromosome Y, mitochondrial genes, and level 3 annotations as defined by Gencode v19^73^ were removed from further analysis^47^. Keeping only protein-coding and lincRNA, the final number of genes used was 16,209 for our 3,027 samples. For all 3,027 individuals, complete genotype and transcriptome data were available.

Plasma proteins were measured using Olink® panels known as: Cardiometabolic (CAM), Cardiovascular II (CVDII), Cardiovascular III (CVDIII), Development (DEV), and Metabolism (MET) (Olink Proteomics AB, Uppsala, Sweden) according to the manufacturer’s instructions^74^. Sample QC was performed using internal and external controls^47^. We removed proteins with intensities below level of detection (LOD) in more than 50% of samples. In addition, we excluded four samples missing plate processing information. The remaining missing protein intensities were imputed using the mean from all samples and subsequently all data were rank normalised. The final proteomic data analysed consisted of 373 proteins measured in 3,023 individuals.

Plasma metabolites were assessed using two platforms: (i) 163 metabolites were measured with a FIA-ESI-MS/MS-based targeted metabolomics method, with the AbsoluteIDQTM p150 kit (BIOCRATES Life Sciences AG, Innsbruck, Austria). Quality assessment evaluated peak shapes, retention times, compound identity, and the number of samples with zero values in the metabolites concentration, removing any individual with more than 50% of zeros. Metabolites with concentration below the LOD were discarded^47^; (ii) using an untargeted approach, metabolite concentrations were measured using Liquid Chromatography-Tandem Mass Spectrometry (LC-MS/MS). Raw data were extracted, peak-identified and QC processed using Metabolon’s hardware and software^47^. The final metabolomic datasets analysed consisted of 116 metabolites measured for 3,027 individuals in the targeted approach and 233 metabolites measured for 2,996 individuals for the untargeted approach.

### T2D status associations with molecular phenotypes

An initial linear regression analysis on rank normalised gene expression, protein and metabolite levels was performed to identify molecular phenotypes associated with T2D status, accounting for the additive effects of age, sex and BMI. In addition, Working Package (WP) was included as a covariate in our models to account for the different processing of the samples in the two cohorts (WP2.1 pre-diabetic and WP2.2 diabetic status) along with other technical variables specific to gene expression. T2D status overlapped with the cohort membership (WP2.1 and WP2.2) for all but 105 of the participants, and sample processing between cohorts was also highly correlated with T2D status (ρ = 0.92; p < 2.2×10^−308^), which would explain the lack of significant associations. To assess this, we also performed T2D associations without accounting for the effects of different processing between Working Packages, a variable correlated with T2D status, and found 26 significant associations out of 16,209 genes and again limited protein and metabolite associations, indicating limited power to detect associations with disease status. A WP membership variable was nonetheless included in all further analyses as it described differences in data collection.

Models were fitted using stats::lm() in R version 4.2.0^75^ as follows:

For gene expression, we used a linear model with the following fixed variables: *Gene Expression ∼ Age + Sex + BMI + T2D + WP + Centre + GC Mean Content + Insert Size + Rin Score + Date of Sequencing*; *Gene Expression ∼ Age + Sex + BMI + T2D + Centre + GC Mean Content + Insert Size + Rin Score + Date of Sequencing*. Technical variables included: (i) an indicator for each of the 7 recruitment centres (Centre); (ii) the mean of the percentage of guanine and cytosine content across samples per mRNA molecules (GC Mean Content); (iii) the length of the mRNA molecules (Insert Size); (iv) the RNA integrity number (RIN) and (v) the date of sequencing as a categorical variable.

For proteins, the model included the following variables: *Protein Levels ∼ Age + Sex + BMI + T2D + WP + Centre + Protein Plate*. Protein plate refers to each of the five protein panels evaluated (CAM, CVDII, CVDIII, DEV and MET) and was included to account for any variability due to sample distribution across plates. Since the date of processing for the protein measurements was confounded with the protein plate, we did not include any date information in the proteomics model.

For targeted metabolites, the model included the following variables: *Metabolite Levels ∼ Age + Sex + BMI + T2D + WP + Centre + Metabolite Plate + Date of Processing*. Metabolite plate was the equivalent of protein plate, included to account for the experimental procedure. For untargeted metabolites, we did not include metabolite plate, as it did not apply.

The binary sex variable was represented with 1 for male and 0 for female, giving a positive beta coefficient (β_sex_ > 0) for a sex association with a molecular phenotype when its levels were higher in men compared to women, independently of age, BMI and HbA1c. Multiple testing correction for each variable and each of the four molecular phenotype sets was assessed independently using the Storey and Tibshirani q-value method implemented in the q-value R package^46,76^. Associations with an adjusted p-value ≤ 0.05 after controlling for the False Discovery Rate (FDR) were considered significant.

### Differential analysis of molecular phenotype levels

Associations with gene expression, protein and metabolite levels were examined using linear regression models on rank normalised data, including technical variables specific to each molecular phenotype. Models were fitted using stats::lm() in R version 4.2.0^75^. More specifically:

For gene expression, we used a linear model with the following fixed variables: *Gene Expression ∼ Age + Sex + BMI + HbA1c + WP + Centre + GC Mean Content + Insert Size + Rin Score + Date of Sequencing*.

For proteins, the model included the following variables: *Protein Levels ∼ Age + Sex + BMI + HbA1c + WP + Centre + Protein Plate*.

For targeted metabolites, the model included the following variables: *Metabolite Levels ∼ Age + Sex + BMI + HbA1c + WP + Centre + Metabolite Plate + Date of Processing*. For untargeted metabolites, we did not include metabolite plate, as it did not apply.

### Functional enrichment analyses

To explore which biological processes related to gene expression were enriched across associated genes, we performed gene set enrichment analyses (GSEA) in R using clusterProfiler^77^ with functional annotations from Gene Ontology^78^ (GO), using the entire genome as background. GO biological process terms were summarised using REVIGO^79^ (which allows to backtrace the enriched GO terms to their parent nodes. Subsequently, we calculated the frequency of appearance of these parent nodes of significant GO terms.

For proteins significantly associated with age, sex, BMI and HbA1c we performed enrichment analyses on the Protein-Protein Interaction (PPI) networks using STRING database^80,81^. We evaluated enrichments against the 373 tested proteins as background, as well as against the whole genome. All PPI networks (SFig.27-30) were constructed using protein interactions derived from experiments and databases with the highest confidence interaction score (0.9). Finally, we used REVIGO^79^ as described above.

To investigate the biological significance of associated metabolites we considered metabolites from the targeted and untargeted methods together. We only considered metabolites that corresponded to a KEGG ID (80, 85, 82 and 26 metabolites for age, sex, BMI and HbA1c respectively) in order to perform an enrichment analyses using the R package FELLA^82^. We conducted a diffusion enrichment using the normal approximation to calculate enrichment scores (p.scores). This method involves a sub-network analysis that utilises contextual data from the KEGG database, thus inferring the enriched KEGG pathways. In addition, we constructed the corresponding networks that involved modules, enzymes and reactions that link input compounds associated with age, sex, BMI and HbA1c to their related pathways.

### Interaction analyses between metabolic disease-relevant traits

To examine context-dependent effects between metabolic disease-relevant traits, we implemented interaction models that included age-by-sex (Age*Sex), age-by-BMI (Age*BMI) and age-by-HbA1c (Age*HbA1c) interaction terms for each molecular phenotype dataset. We residualised gene expression, protein and metabolite levels in order to remove the effect of biological covariates that were not to be included in the interaction model as well as the effect of technical covariates per molecular phenotype. Before implementing the models, numerical variables to be included in each interaction model were centred to the mean in order to account for the mediating non-essential multicollinearity and so that main effect terms and the interaction term could be comparable^83–85^. Molecular phenotypes were tested for context-dependent effects of age with the following linear interaction models fitted using stats::lm() in R:

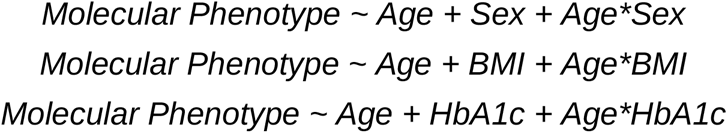

We examined sex-specific effects of metabolic markers by implementing interaction models that included BMI-by-sex (BMI*Sex) and HbA1c-by-sex (HbA1c*Sex) interaction terms for each molecular phenotype dataset. We also examined BMI-dependent effects of HbA1c levels. Molecular phenotypes were tested for these context-dependent effects of metabolic markers using the following interaction models:

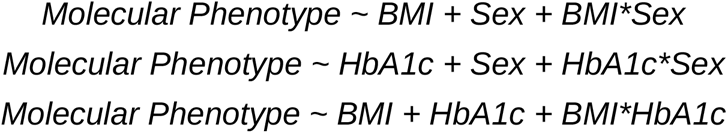

Multiple testing correction for each molecular phenotype type was assessed independently using the q-value method implemented in the q-value R package. Molecular phenotypes significant for context-dependent effects of age were those with an adjusted p-value ≤ 0.05 after controlling for the FDR.

For visualisation purposes, stratified analyses were performed; For sex, we performed a stratified linear regression separately for male and female individuals using the same models as before to calculate the rate of change with age, BMI or HbA1c within each sex by performing a t-test^86^ using stats::t.test() in R (t-test p ≤ 0.05). Differences in rate of change were determined by comparing the β coefficients calculated separately for male and female individuals while the direction of effect was assessed by the sign of the β in each sex. For BMI, linear regression calculations were performed separately for individuals within the BMI classification groups: normal, overweight and obese individuals. We used the World Health Organisation (WHO) standards and definitions as: Underweight < 18.5, 18.5 ≤ Normal < 25, 25 ≤ Overweight < 30, Obese ≥ 30. We did not attempt to fit a linear regression model for underweight individuals as there were only 3 individuals in the cohort. The difference in rate of change with age, BMI or HbA1c was determined by comparing the β coefficients for individuals with normal, overweight and obese BMI while the direction of effect was assessed by the sign of the βage in each BMI group. For HbA1c-dependent age effects, we calculated the rate of change with age (β_age_) between individuals with low (20% of individuals with the lowest HbA1c) and high (20% of individuals with the highest HbA1c) levels of HbA1c. Linear regression calculations were performed separately for each group as described previously.

### Relationships between molecular phenotypes

To explore associations between pairs of molecular phenotypes we used linear regression models with residuals from gene expression, protein and metabolite levels after removing the effect of age, sex, BMI, HbA1c and technical covariates specific per molecular phenotype. We explored relationships across different types of molecules. All five possible pair-wise comparisons between molecular phenotypes were evaluated using stats::lm() in R as follows:

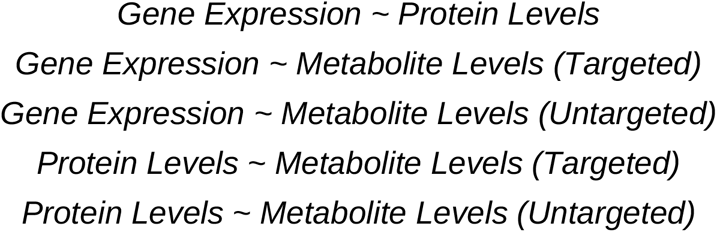

Multiple testing correction was assessed for each pair-wise comparison model using the q-value method to control the FDR, with an adjusted p-value ≤ 0.05.

### Context-dependent relationships between pairs of molecular phenotypes

We investigated relationships between pairs of molecular phenotypes where levels of one phenotype affected the levels of another depending on age, sex, BMI or HbA1c levels. We used interaction models, per clinical variable, for all pair combinations across molecular phenotype sets (gene expression, protein and metabolite levels). Residuals of gene expression, protein and metabolite levels were used after removing the effect of technical covariates per molecular phenotype. For each studied trait, the remaining biological covariates that were not included in the interaction term, were included in the the model:

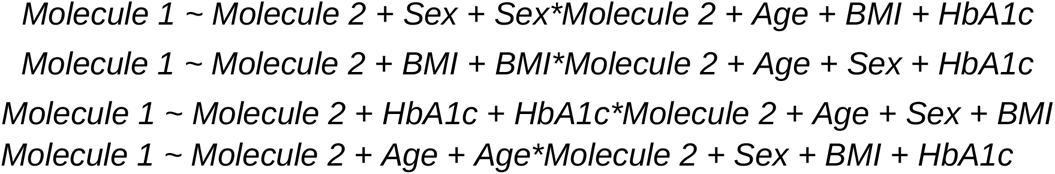

Pairs of molecular phenotypes were tested using stats::lm() in R. Multiple testing correction was assessed for each model using the q-value method to control the FDR, with an adjusted p-value ≤ 0.05.

### Direction of causality between age-related molecular phenotypes

To determine the causal relationships between age and pairs of associated molecular phenotype pairs we implemented a Bayesian network analysis^87^. We first identified all associated pairs of age-related molecular phenotypes, including pairs within the same molecule type. Subsequently, we considered network topologies that assume a causal effect from age towards molecules, as the opposite effect does not have biological meaning since the age of an individual increases regardless of the level of any molecular phenotype. Three models were evaluated (SFig.26c): I) age affects molecule 1 which in turn affects molecule 2; II) age affects molecule 2, which in turn affects molecule 1; III) age affects both molecules independently. The probabilities can be described with the following formulas:

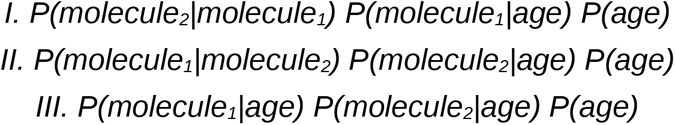

Only pairs of molecular phenotypes where both phenotypes were previously associated with age were tested. We used residuals for gene expression, protein and metabolite level after removing the effect of sex, BMI, HbA1c and technical covariates specific per molecular phenotype to compute the likelihood of the three possible Bayesian network topologies using the bnlearn R package^88,89^. The best fit across the three models was evaluated using the Bayesian Information Criterion (BIC) score: for every age-molecule pair trio, if one of the three models had a BIC score > 10 compared to each of the other two models, we considered that there was very strong evidence supporting that model. Similarly, a BIC score difference ≥ 6 and ≤ 10 indicated strong evidence. For BIC score differences < 6 the trio was not considered for further evaluation.

### Network of age-related molecular phenotypes and genetic associations

Using Cytoscape v3.10.1 we constructed a network of molecular phenotypes significantly associated with age (S78). Edges indicated significant associations between pairs of age-related molecules including the nominal p-value of the associations as weight. Direction of edges was assigned using the causal inference results (BIC score>= 6). Edges between pairs of molecules independently affected by age were erased in order to show only directions from dependent models. We also included SNPs as nodes, with arrows indicating their influence on the levels of the age-related molecules, from Quantitative Trait Loci (QTLs) identified by Brown et al.^47^ in the DIRECT dataset. The network initially included 11,169 molecules and 10,228 SNPs as nodes and >9.8×10^7^ edges connecting them, showing significantly associated pairs of age-related molecules or QTLs. For visualisation purposes we omitted the 9.7×10^7^ associations between gene expression pairs, resulting in a network of 11,166 molecules and 10,226 SNPs as nodes and ∼1.1×10^6^ edges representing either significant associations between age-related molecule pairs or QTLs.

To visualise a subset of the mTOR signalling pathway, we selected genes and proteins that are part of the pathway in our age network along with their connections, omitting associated pairs of gene expressions. We used the gene set provided by GSEA^90,91^ to determine the mTOR signalling pathway gene set (52 genes), as contributed by KEGG^92–94^. In our age network, 35 nodes were part of the mTOR gene set and the corresponding proteins. For visualisation purposes, we selected the 1^st^ neighbours of these nodes (directly associated molecules; FDR ≤ 0.05) and only considered the edges of strongly associated pairs of molecules (FDR ≤ 10^−10^). Genetic effects were omitted for this network. Directionality of edges between molecular phenotype pairs was defined by the Bayesian network results, meaning that the effect of age was mediated by the source molecule towards the target molecule. We removed edges with no BIC score above 10 and edges that best fit the independent model. Finally, again for visualisation purposes, proteins with degree<=3 and metabolites with degree=1 were removed, resulting in a network of 74 nodes and 238 edges. Subsequently, we highlighted the molecules/nodes that were also independently associated with sex depending on whether their levels were higher or lower in men compared to women.

To visualise a T2D-related pathway, we focused on a subset of our directed age network around molecules/nodes found to be influenced by a SNP (previously identified QTLs in the DIRECT dataset^47^) found to be colocalised with a T2D GWAS^95^. For visualisation purposes, we selected the 1st neighbours of these nodes (directly associated molecules; FDR ≤ 0.05) and considered edges of associated pairs of molecules with FDR ≤ 0.001). Directionality of edges between molecular phenotype pairs was defined by the Bayesian network results, removing edges with no BIC score and edges that best fit the independent model. Finally, proteins with degree<90 and metabolites with degree<65 were removed, resulting in a network of 67 nodes and 1,211 edges.

### Colocalisation analyses

To perform colocalisation we used the R package coloc v5.2.3^96^ with SNPs previously identified QTLs in the DIRECT dataset^47^ that influenced levels of age-related molecular phenotypes and SNPs from a T2D GWAS study (Mahajan et al., 2018), before and after controlling for BMI. For every SNP around a 20kpb window of the primary SNP involved in the QTL also measured in the GWAS, two Bayes factors were calculated using the summary statistics from the two analyses. The Bayes factor provides an alternative to the p-value for the ranking of associations and measures relative support for a model in which the SNP is associated with the trait compared to the null model of no association. Evidence for colocalisation between the two traits is considered based on the significance of the H4 posterior probability, indicating association with trait 1 and trait 2 are sharing a SNP.

## Ethics statements

Ethics committee/IRB of Lund University, Regional Ethical Review Board in Lund, Sweden (20130312105459927); Regional Committee on Health Research Ethics for the Capital Region of Denmark, Copenhagen, Denmark (H-1-2012-166 and H-1-2012-100); Medical Ethics Committee of the Academic Medical Center, Amsterdam, Netherlands (NL40099.029.12); and NRES Committee North East - Newcastle North, Dundee and Exeter, UK (12/NE/0132) gave ethical approval for this work. All participants provided written informed consent at enrolment. The research conformed to the ethical principles for medical research involving human participants outlined in the Declaration of Helsinki.

## Data availability

The molecular and clinical raw data as well as the processed data are available under restricted access due to the informed consent given by study participants. The various national ethical approvals for the present study, and the European General Data Protection Regulation (GDPR) impose that individual-level clinical and molecular data cannot be transferred from the centralized IMI-DIRECT repository. Requests for access will be informed on how data can be accessed via the DIRECT secure analysis platform following submission of an appropriate application. The IMI-DIRECT data access policy is available at https://directdiabetes.org.

Code for data analysis is available on https://github.com/dafniLettos/DIRECT_metabolic

Supplementary data and results tables can be found at https://doi.org/10.5281/zenodo.18281553

## Acknowledgements

The work leading to this publication has received support from the Innovative Medicines Initiative Joint. Undertaking under grant agreement n°115317 (DIRECT, https://directdiabetes.org/), resources of which are composed of financial contributions from the European Union Seventh Framework Programme (FP7/2007-2013) and EFPIA companies’ in-kind contribution. We thank all the participants and study centre staff in IMI DIRECT for their contribution to the study.

