## Supplemental Figures for "Context-dependent molecular responses to heterogeneous metabolic disease traits" (from another version of this preprint: version 1, 2026-06-08)

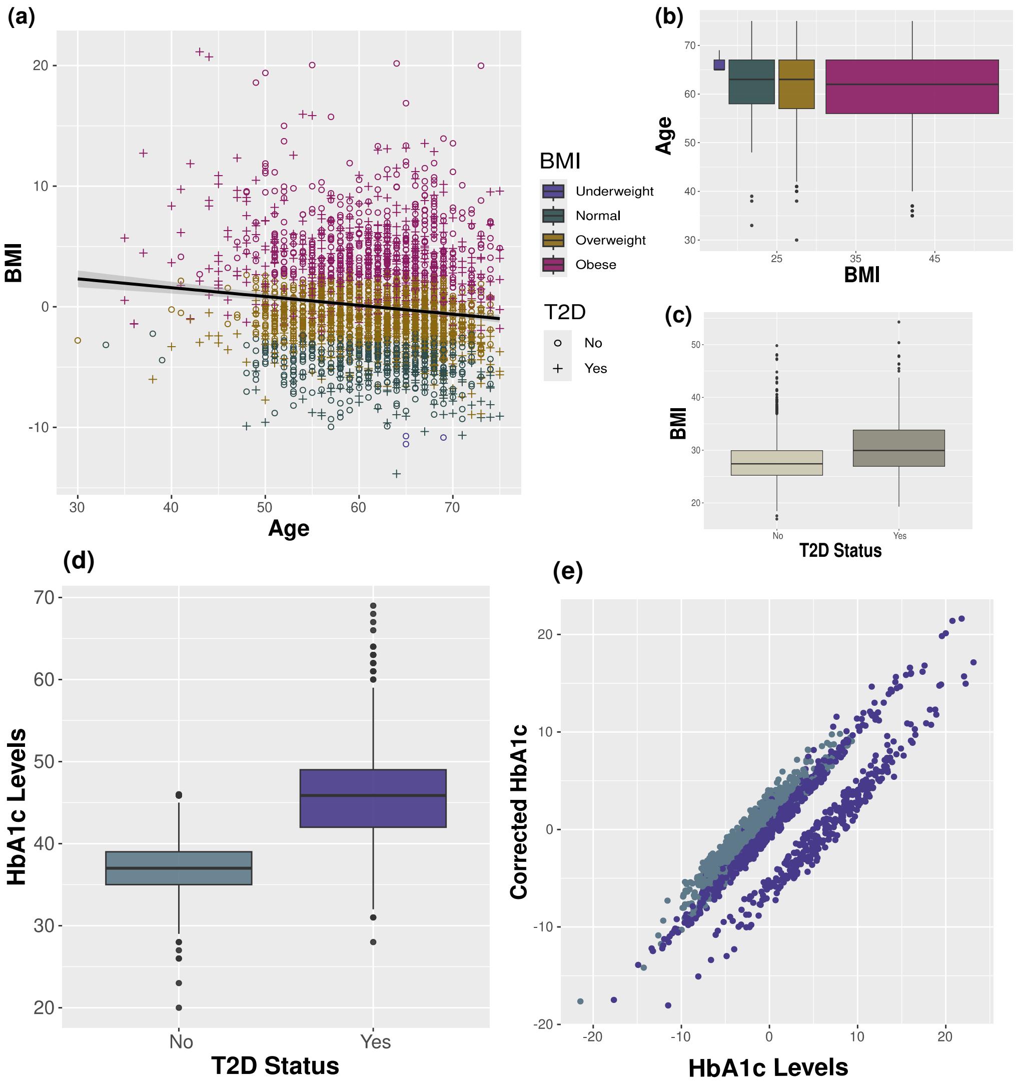


Sup. Figure 1: Relationship between variables and clinical traits in the DIRECT cohort. (a) Relationship between age and BMI; Colours indicate BMI classification according to the World Health Organisation (WHO) standards: Underweight<18.5, 18.5≤Normal<25, 25≤Overweight<30, Obese≥30. Shape indicates disease status for T2D for pre-diabetic (No) and diabetic individuals (Yes); b. Distribution of individuals across BMI bands and age; c. Distribution of BMI for pre-diabetic and diabetic individuals. d. Distribution of fasting HbA1c levels (mmol/mol) for pre-diabetic and diabetic individuals; e. Comparison of HbA1c values before and after accounting for the Working Package (WP) variable. HbA1c is correlated with WP and T2D status ($\rho = 0.66$). However, for diabetic individuals (WP 2.2) especially, accounting for HbA1c would not be equivalent to fully accounting for T2D status as additional criteria was used to assign disease status.

(a)

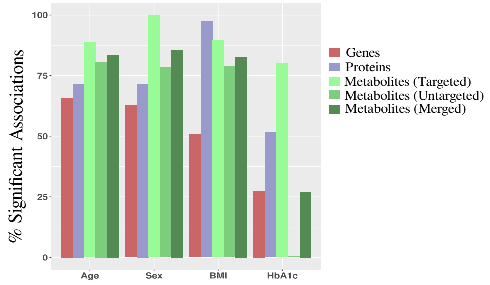


(d)

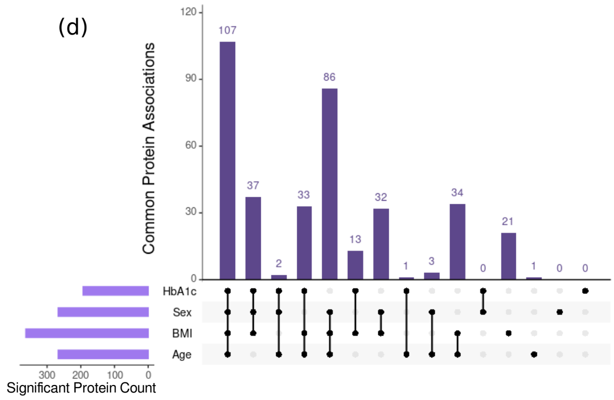


(b)

| | Age Associations | Sex Associations | BMI Associations | HbA1c Associations | Tested |
|---|---|---|---|---|---|
| **Genes** | 10,643 | 10,163 | 8,256 | 4,429 | 16,209 |
| **Proteins** | 267 | 267 | 363 | 193 | 373 |
| **Metabolites (Targeted)** | 103 | 116 | 104 | 93 | 166 |
| **Metabolites (Untargeted)** | 188 | 183 | 184 | 1 | 233 |
| **Metabolites (Merged)** | 291 | 299 | 288 | 94 | 349 |

(c)

(e)

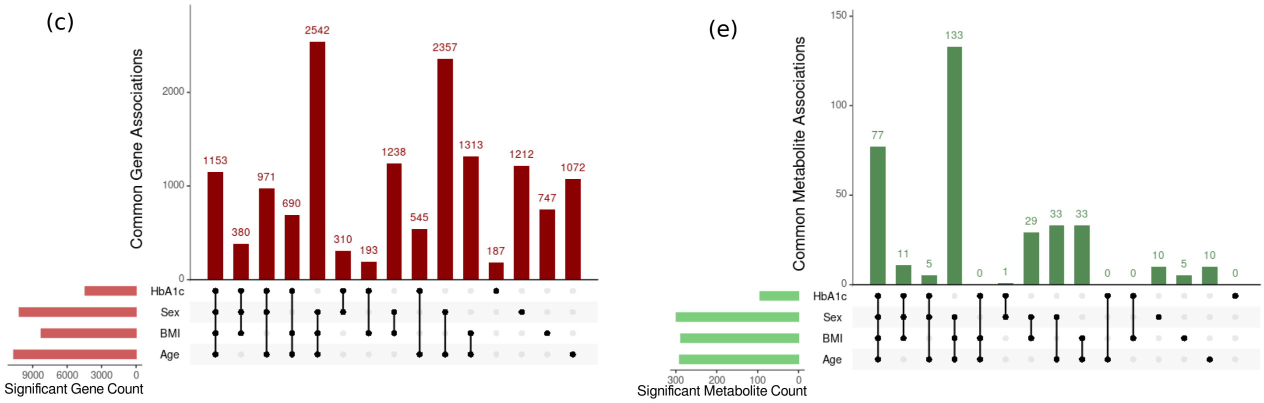


Sup. Figure 2: Molecular phenotypes significantly associated with biological traits. (a) Percentages of molecular phenotypes associated with age, sex, BMI and HbA1c per molecular phenotype dataset; Significance of associations was assessed using linear models and a threshold of FDR-adjusted Pvalue ≤ 0.05. (b) Upset plot showing all intersections of significantly associated genes across all biological traits tested (11.4% overlap across all traits). (c) Upset plot showing all intersections of significantly associated proteins across all biological traits tested (37.5% overlap across all traits). (d) Upset plot showing all intersections of significantly associated metabolites across all biological variables tested (22.1% overlap across all traits).

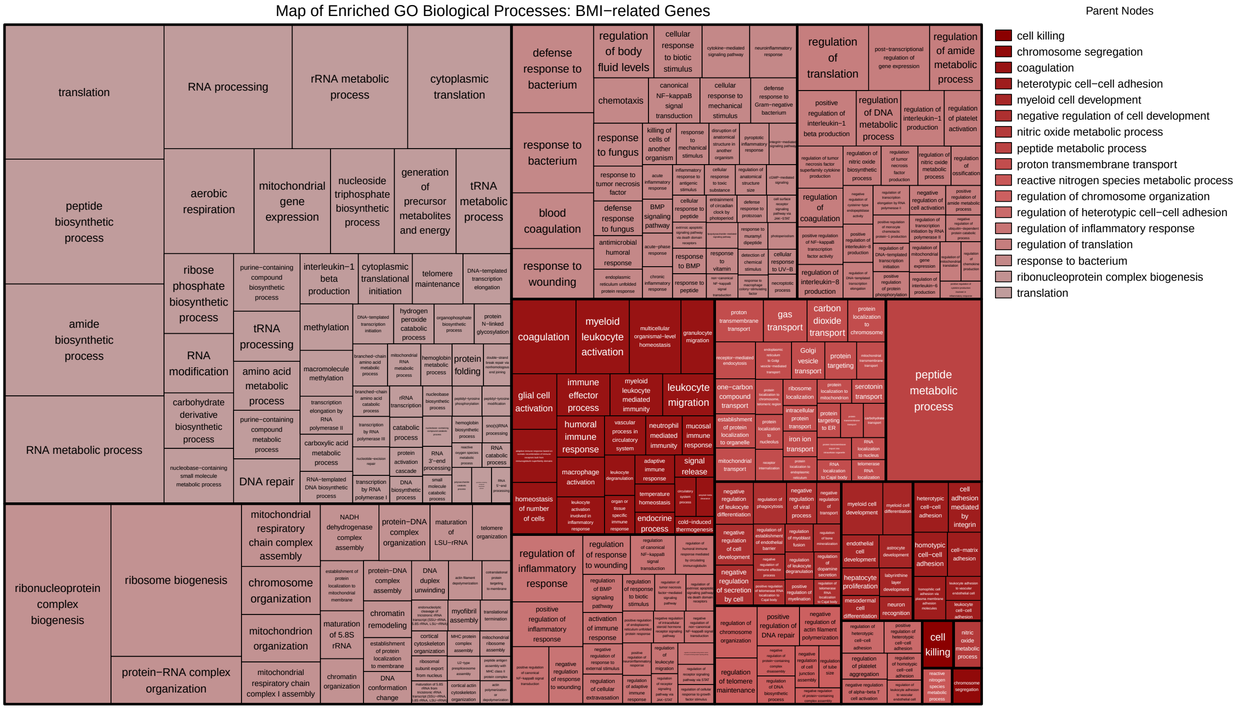


Sup. Figure 3: Treemap representing and summarising the list of Gene Ontology (GO) biological process terms enriched in BMI-related genes. GO terms are displayed as nested rectangles, where the size of each rectangle is proportional to the statistical significance ($\log_{10}$P-value$_{enrichment}$) of the corresponding GO term. Terms are grouped into higher-level categories based on semantic similarity, with each category, referred to as a parent node, represented by a distinct colour.

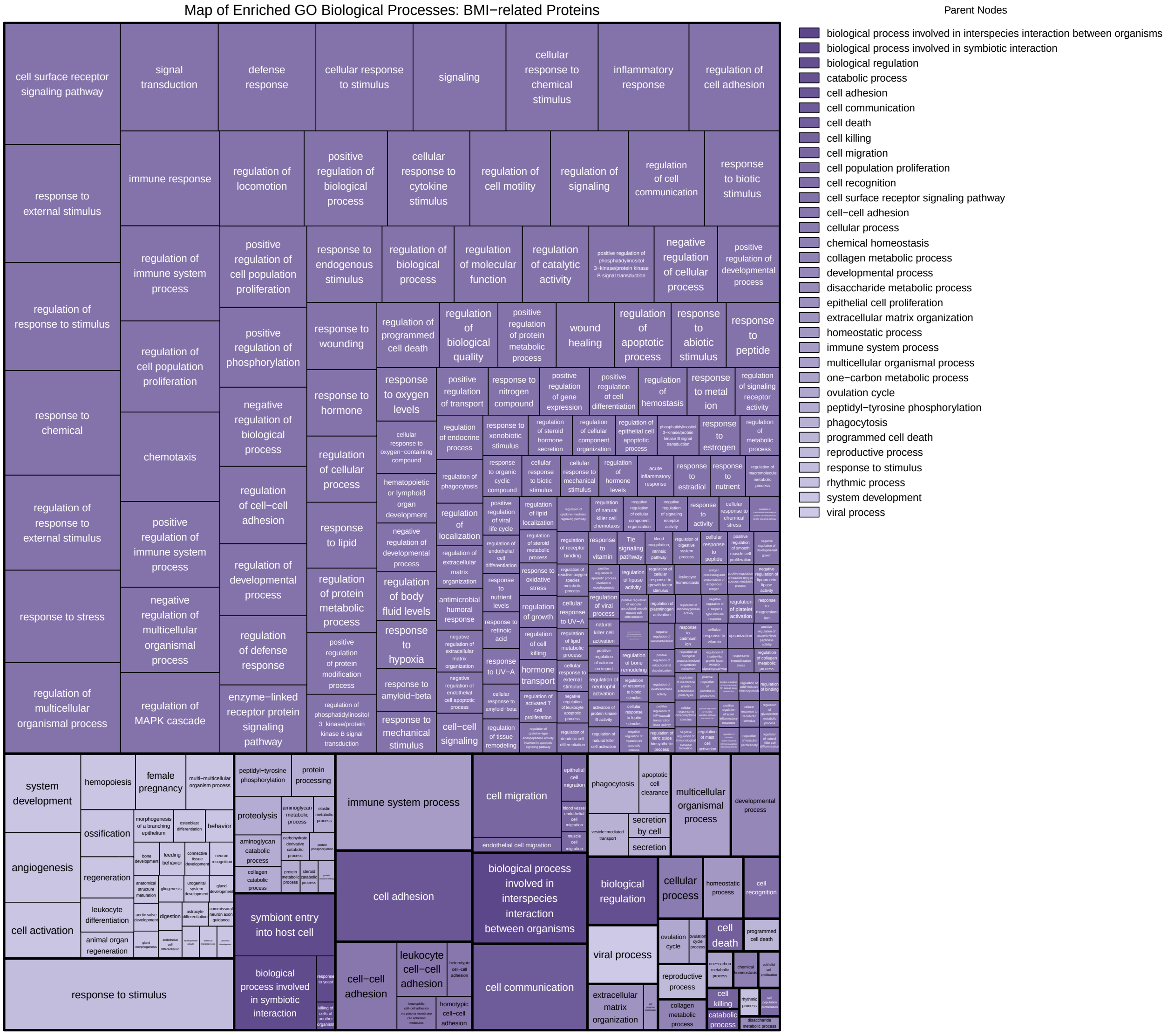


Sup. Figure 4: Treemap representing and summarising the list of Gene Ontology (GO) biological process terms enriched in BMI-related proteins. GO terms are displayed as nested rectangles, where the size of each rectangle is proportional to the statistical significance ($\log_{10}$P-value$_{enrichment}$) of the corresponding GO term. Terms are grouped into higher-level categories based on semantic similarity, with each category, referred to as a parent node, represented by a distinct colour.

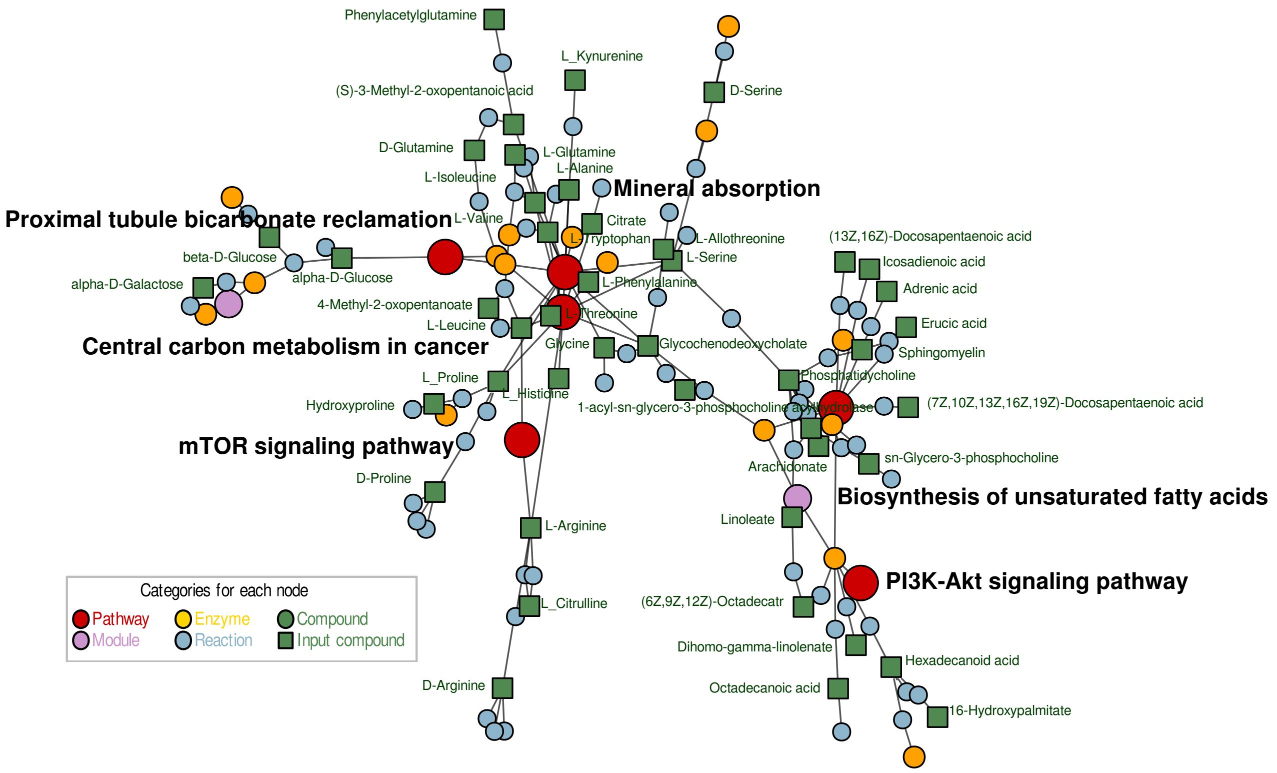


Sup. Figure 5: Network constructed with the diffusion method of the R package FELLA, using contextual data from the KEGG database: green nodes represent BMI-related metabolites while red notes represent the KEGG pathways enriched within the list of BMI-related metabolites.

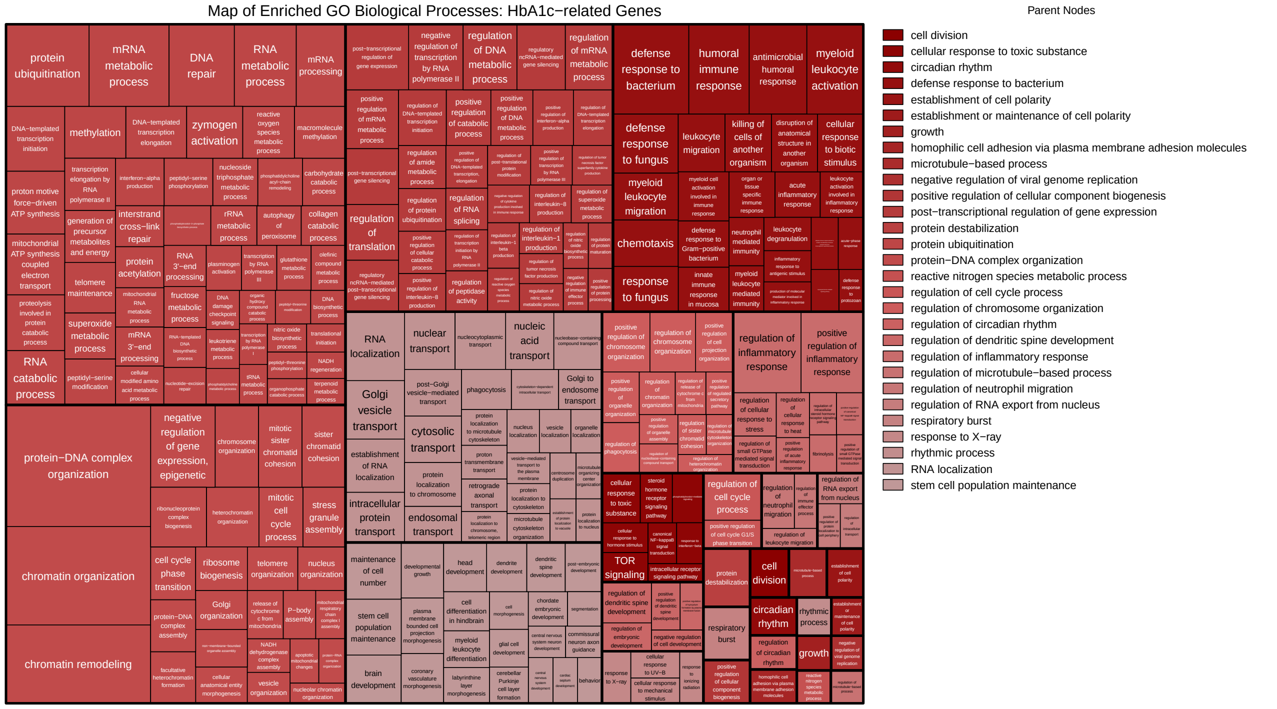


Sup. Figure 6: Treemap representing and summarising the list of Gene Ontology (GO) biological process terms enriched in HbA1c-related genes. GO terms are displayed as nested rectangles, where the size of each rectangle is proportional to the statistical significance ($\log_{10}$P-value$_{enrichment}$) of the corresponding GO term. Terms are grouped into higher-level categories based on semantic similarity, with each category, referred to as a parent node, represented by a distinct colour.

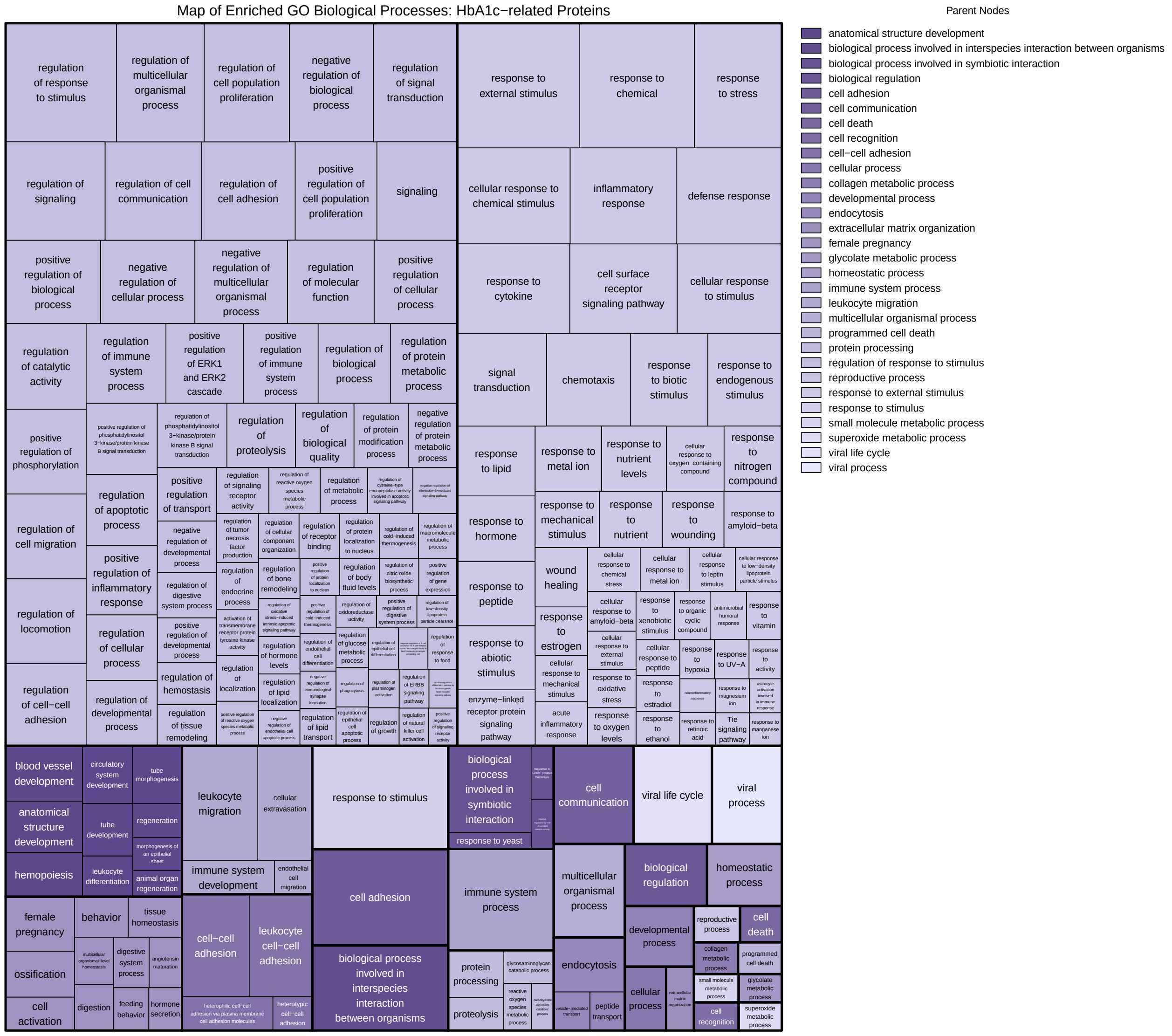


Sup. Figure 7: Treemap representing and summarising the list of Gene Ontology (GO) biological process terms enriched in HbA1c-related proteins. GO terms are displayed as nested rectangles, where the size of each rectangle is proportional to the statistical significance ($\log_{10}$P-value$_{\text{enrichment}}$) of the corresponding GO term. Terms are grouped into higher-level categories based on semantic similarity, with each category, referred to as a parent node, represented by a distinct colour.

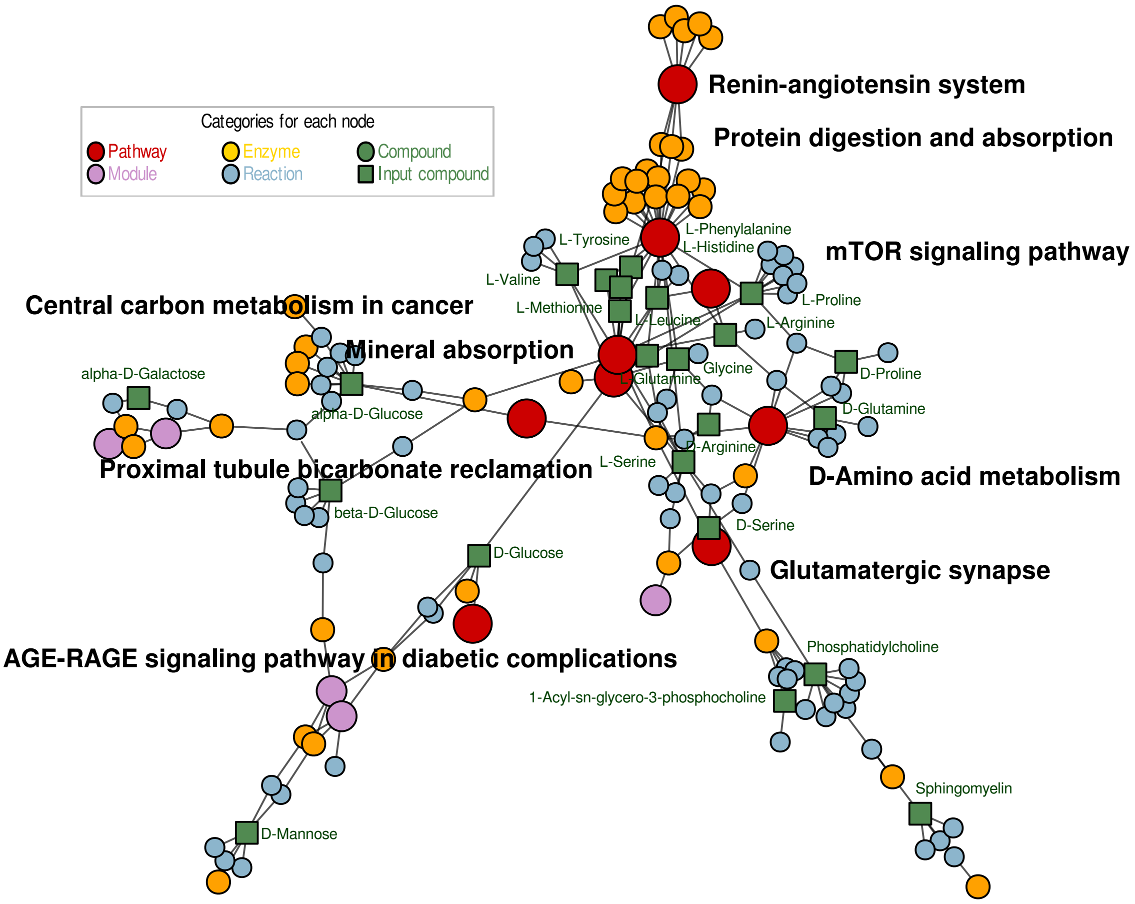


Sup. Figure 8: Network constructed with the diffusion method of the R package FELLA, using contextual data from the KEGG database: green nodes represent HbA1c-related metabolites while red notes represent the KEGG pathways enriched within the list of HbA1c-related metabolites.

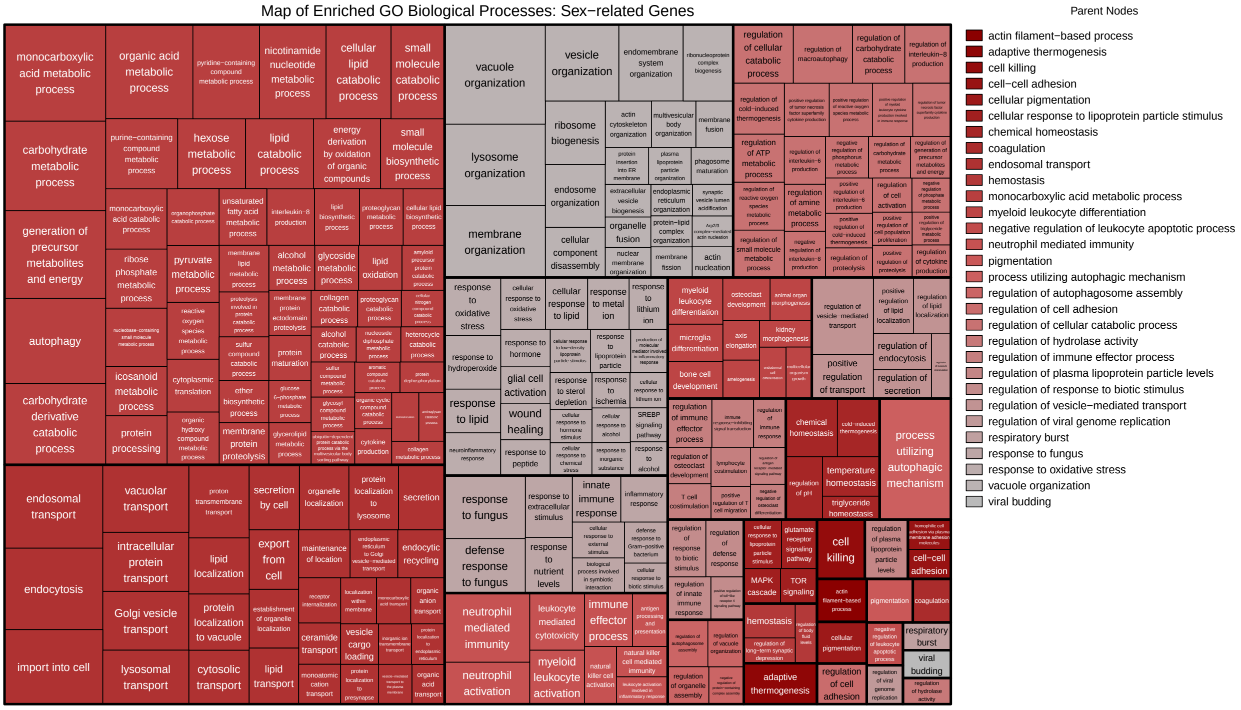


Sup. Figure 9: Treemap representing and summarising the list of Gene Ontology (GO) biological process terms enriched in sex-related genes. GO terms are displayed as nested rectangles, where the size of each rectangle is proportional to the statistical significance ($\log_{10}$P-value$_{enrichment}$) of the corresponding GO term. Terms are grouped into higher-level categories based on semantic similarity, with each category, referred to as a parent node, represented by a distinct colour.

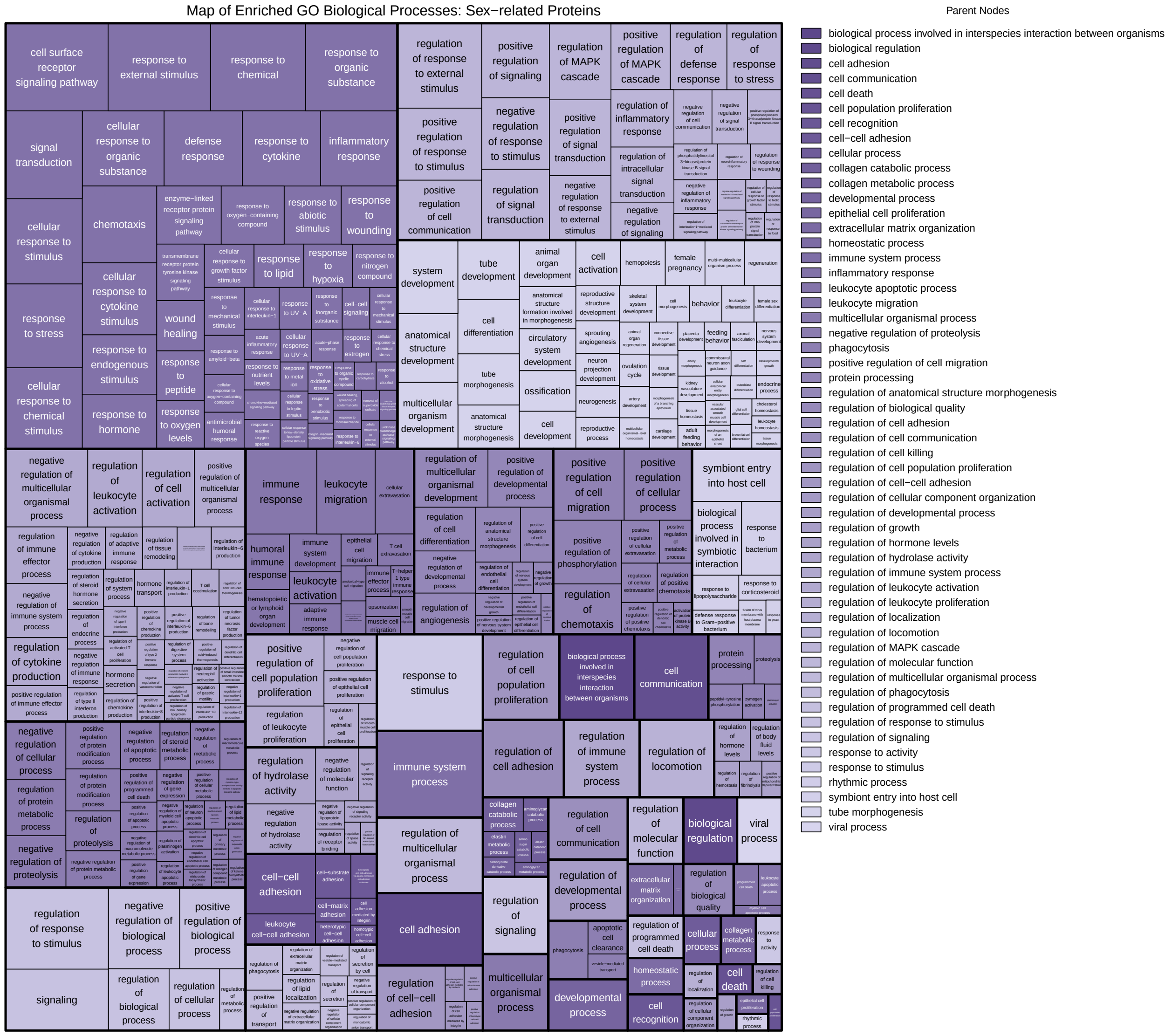


Sup. Figure 10: Treemap representing and summarising the list of Gene Ontology (GO) biological process terms enriched in sex-related proteins. GO terms are displayed as nested rectangles, where the size of each rectangle is proportional to the statistical significance ($\log_{10}$P-value$_{\mathrm{enrichment}}$) of the corresponding GO term. Terms are grouped into higher-level categories based on semantic similarity, with each category, referred to as a parent node, represented by a distinct colour.

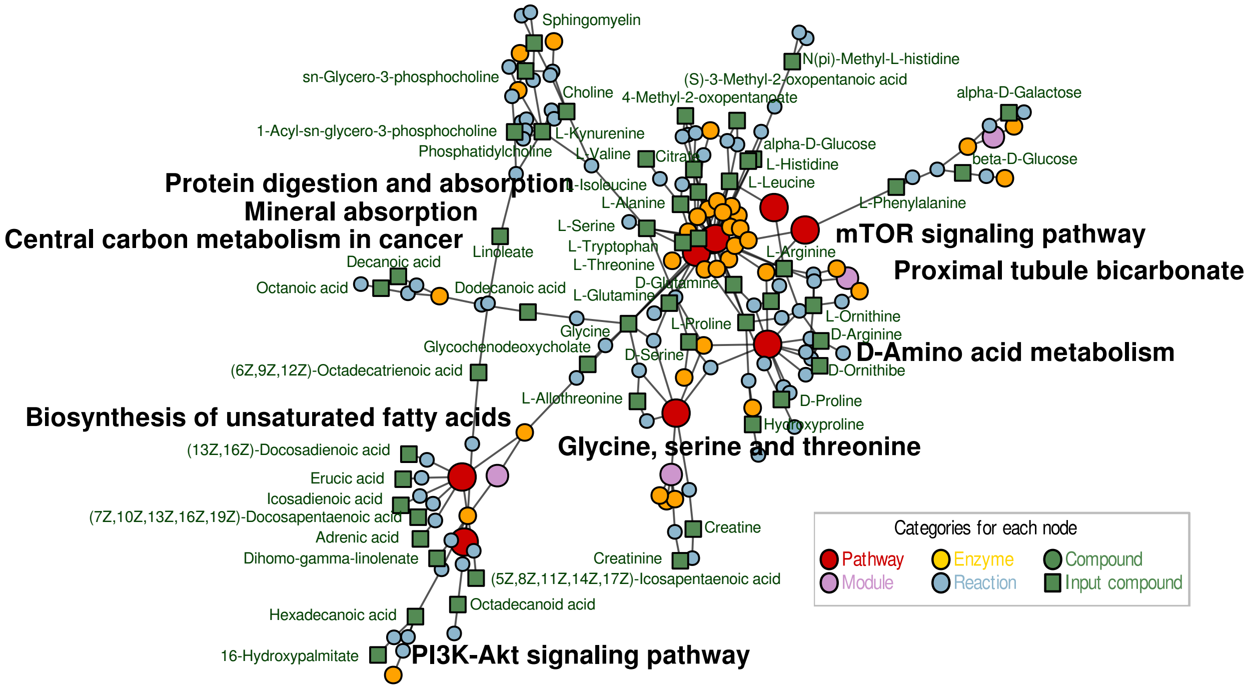


Sup. Figure 11: Network constructed with the diffusion method of the R package FELLA, using contextual data from the KEGG database: green nodes represent sex-related metabolites while red notes represent the KEGG pathways enriched within the list of sex-related metabolites.

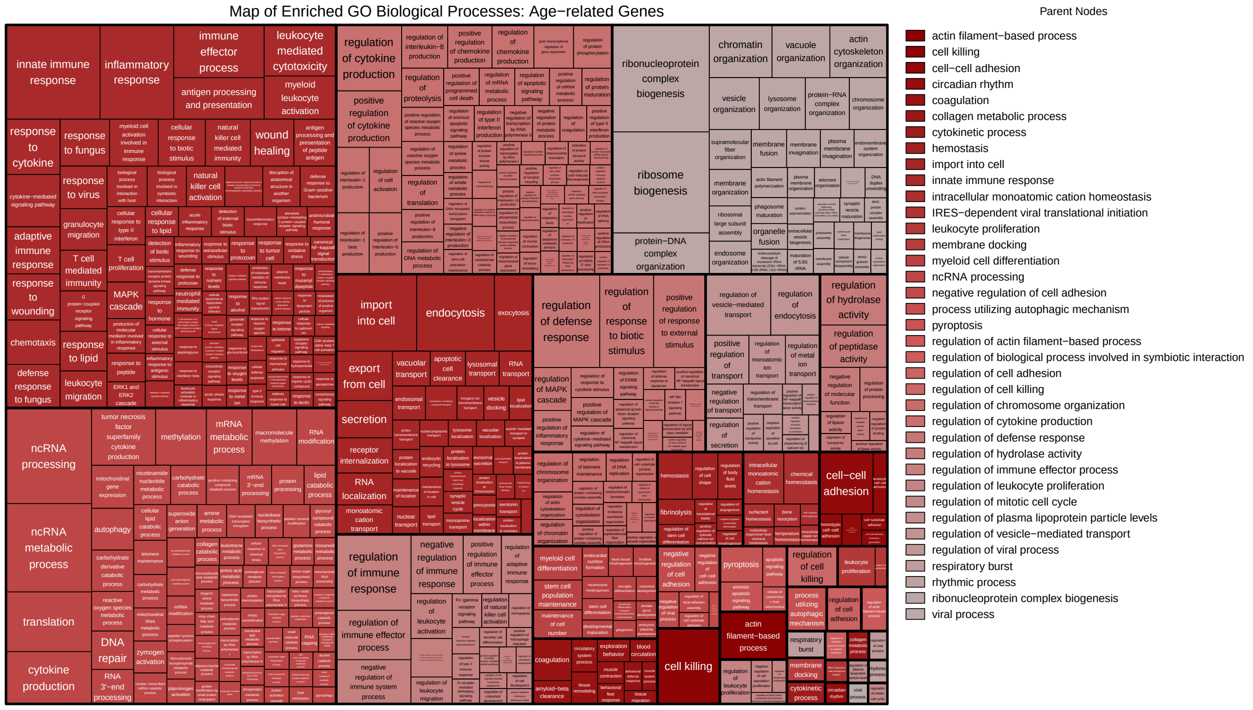


Sup. Figure 12: Treemap representing and summarising the list of Gene Ontology (GO) biological process terms enriched in age-related genes. GO terms are displayed as nested rectangles, where the size of each rectangle is proportional to the statistical significance ($\log_{10}$P-value$_{enrichment}$) of the corresponding GO term. Terms are grouped into higher-level categories based on semantic similarity, with each category, referred to as a parent node, represented by a distinct colour.

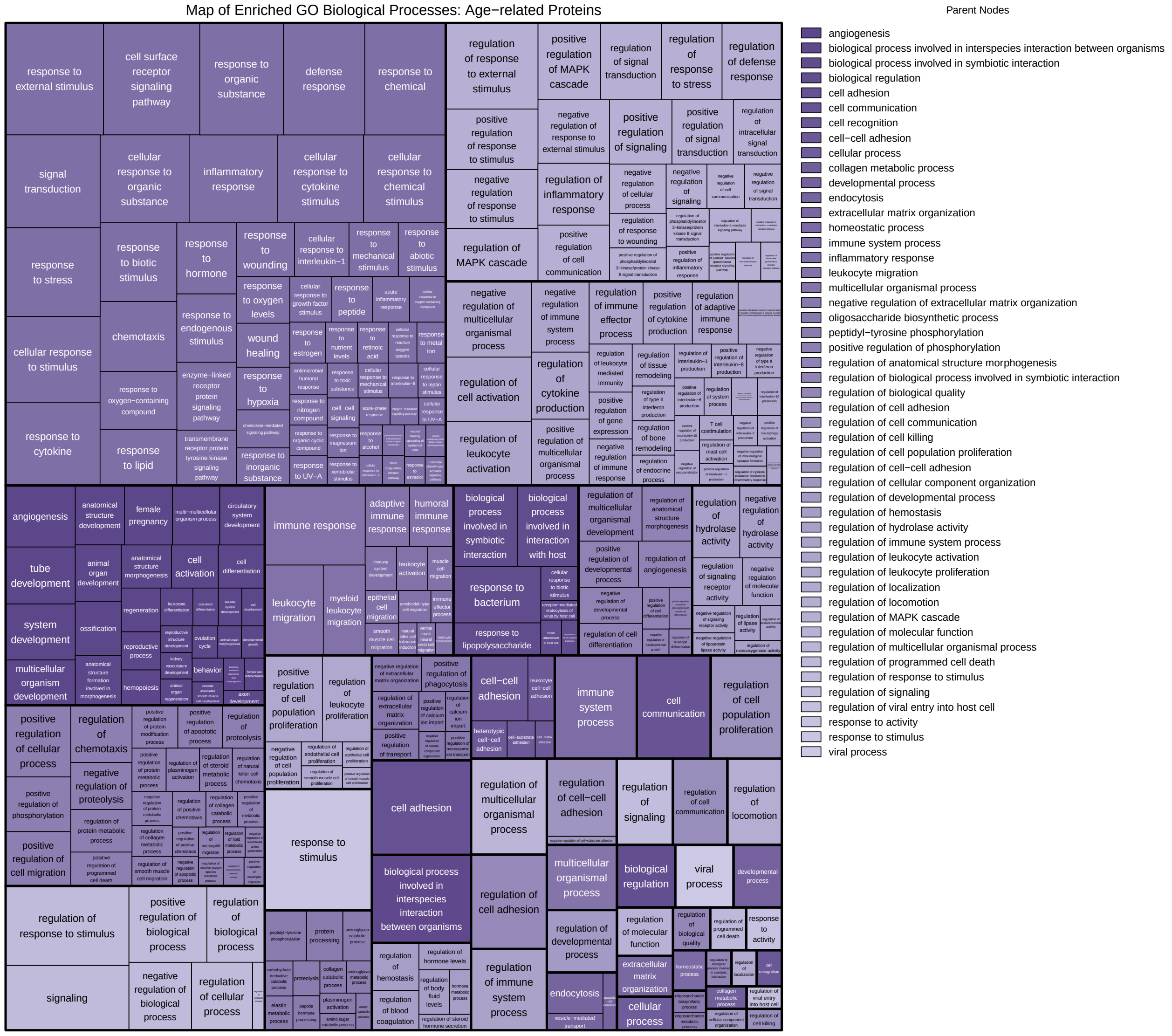


Sup. Figure 13: Treemap representing and summarising the list of Gene Ontology (GO) biological process terms enriched in age-related proteins. GO terms are displayed as nested rectangles, where the size of each rectangle is proportional to the statistical significance ($\log_{10}$P-value$_{enrichment}$) of the corresponding GO term. Terms are grouped into higher-level categories based on semantic similarity, with each category, referred to as a parent node, represented by a distinct colour.

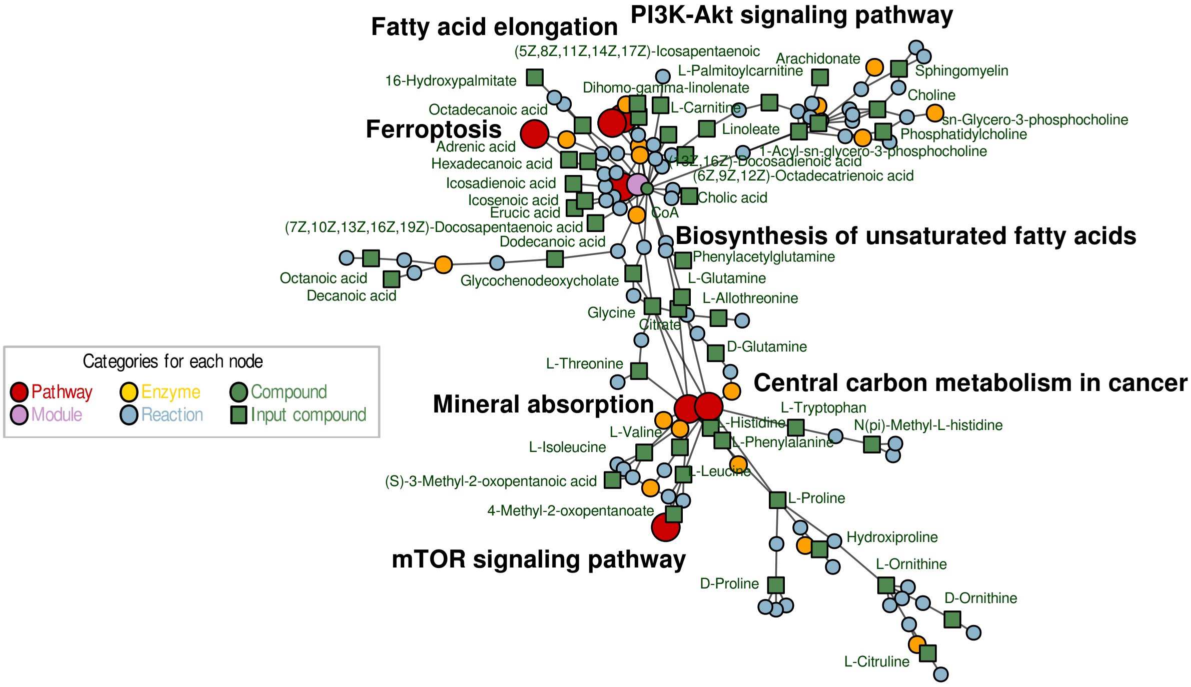


Sup. Figure 14: Network constructed with the diffusion method of the R package FELLA, using contextual data from the KEGG database: green nodes represent age-related metabolites while red notes represent the KEGG pathways enriched within the list of age-related metabolites.

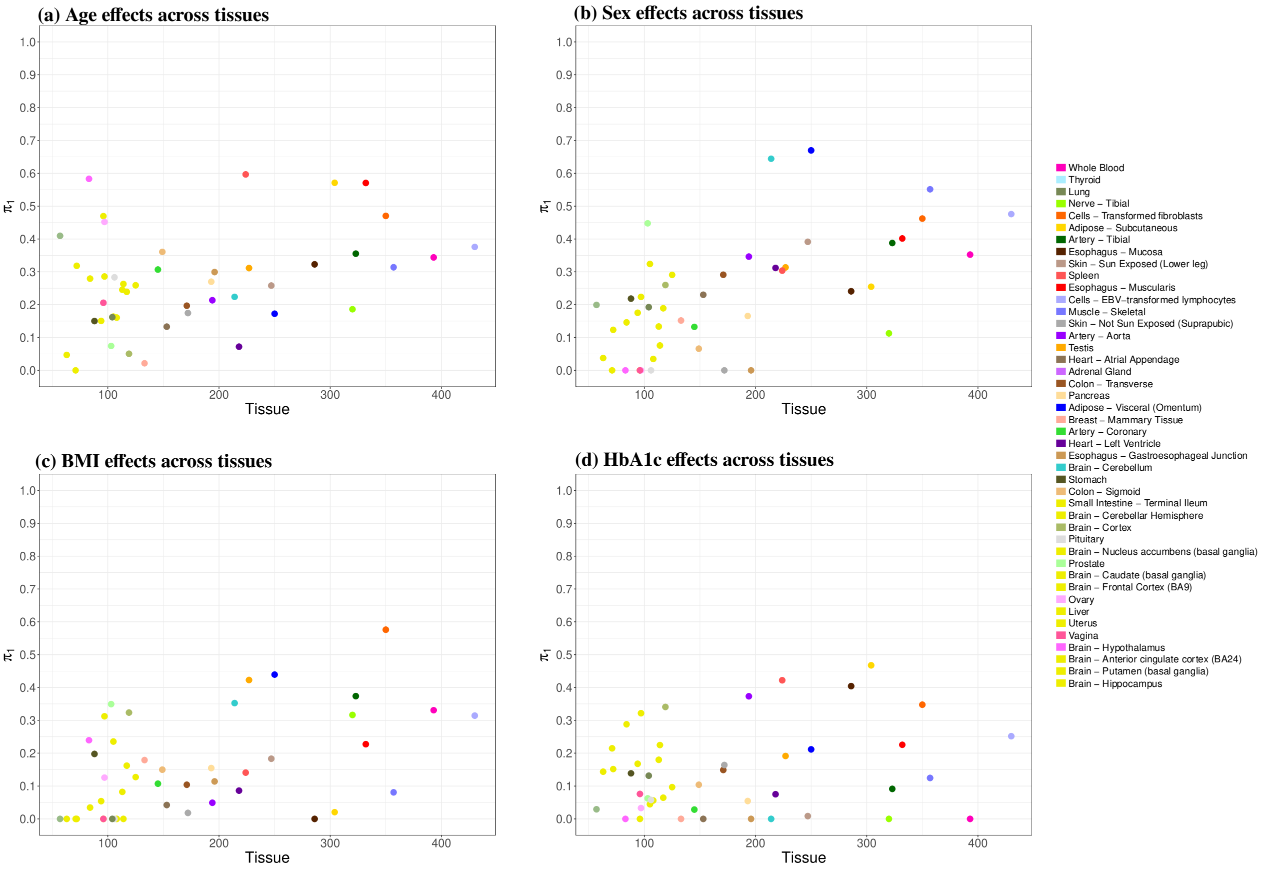


Sup. Figure 15: Comparison of identified gene expression associations with the gene expression multi-tissue GTEx resource using p-value enrichment analysis. Each point represents a tissue coloured according to the GTEx colour. The proportion of true alternative hypotheses (π1) is depicted on the y-axis, while the GTEx tissue sample count is shown on the x-axis for: (a) age associations shared across tissues; (b) sex associations shared across tissues; (c) BMI associations shared across tissues; (d) HbA1c associations shared across tissues. As GTEx does not report associations with HbA1c levels, we performed this comparison with genes significantly associated with T2D status in GTEx since it was a basic criterion for assigning T2D status in the DIRECT cohort and highly correlated with T2D status.

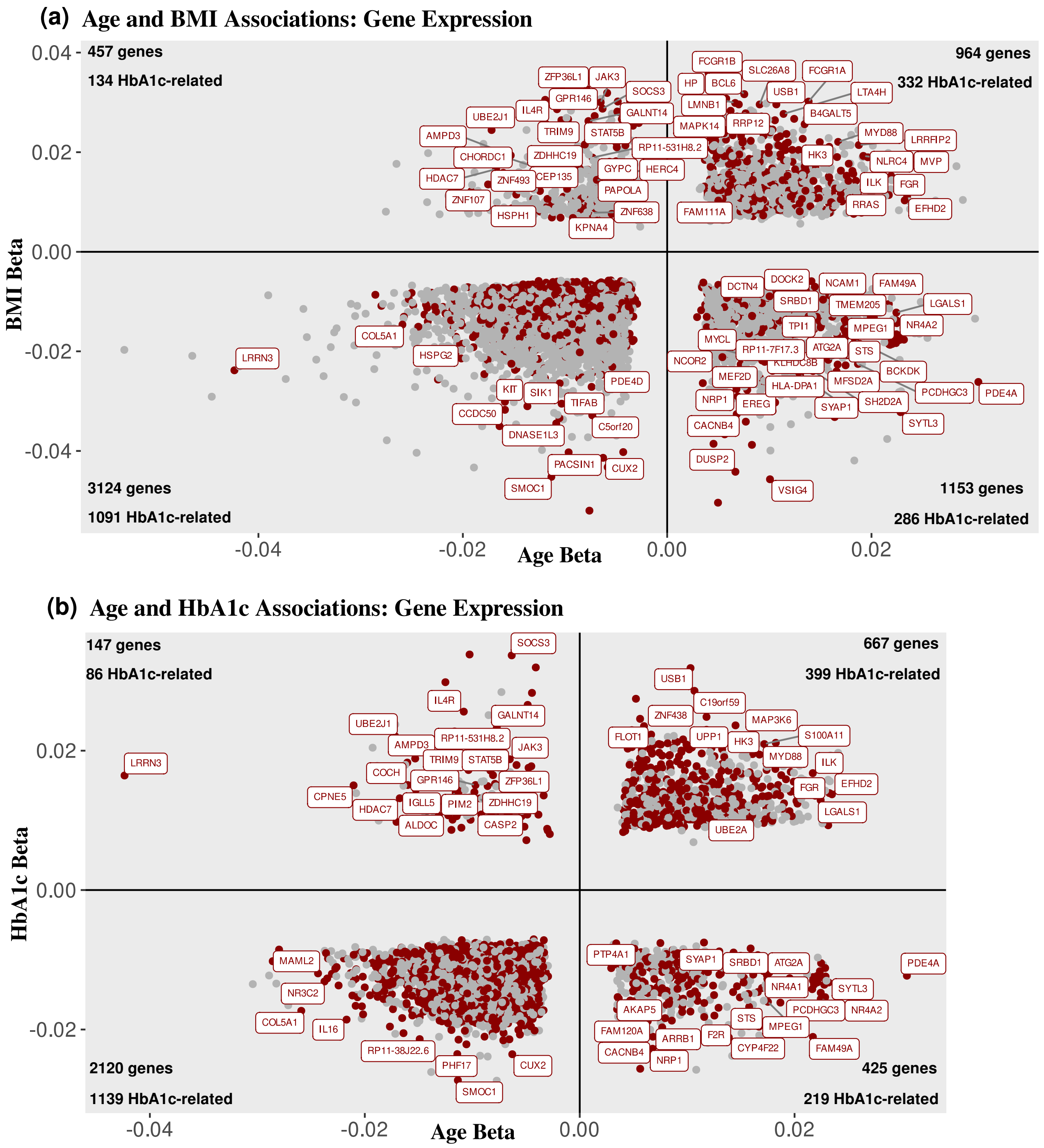


Sup. Figure 16: (a) Genes independently associated with age and BMI (FDR≤0.05). Genes with a positive age beta (right side of y-axis) increase with age while genes with a negative beta (left side of y-axis) decrease with age. Similarly, genes with a positive BMI beta (above x-axis) showed increased expression for higher BMI while genes with a negative BMI beta (below x-axis) showed decreased expression for higher BMI values. Genes in red were also significantly associated with HbA1c. (b) Genes independently associated with age and HbA1c (FDR≤0.05). Genes with a positive age beta (right side of y-axis) increase with age while genes with a negative beta (left side of y-axis) decrease with age. Similarly, genes with a positive HbA1c beta (above x-axis) showed increased expression for higher HbA1c while genes with a negative HbA1c beta (below x-axis) showed decreased expression for higher HbA1c values. Genes in red were also significantly associated with BMI.

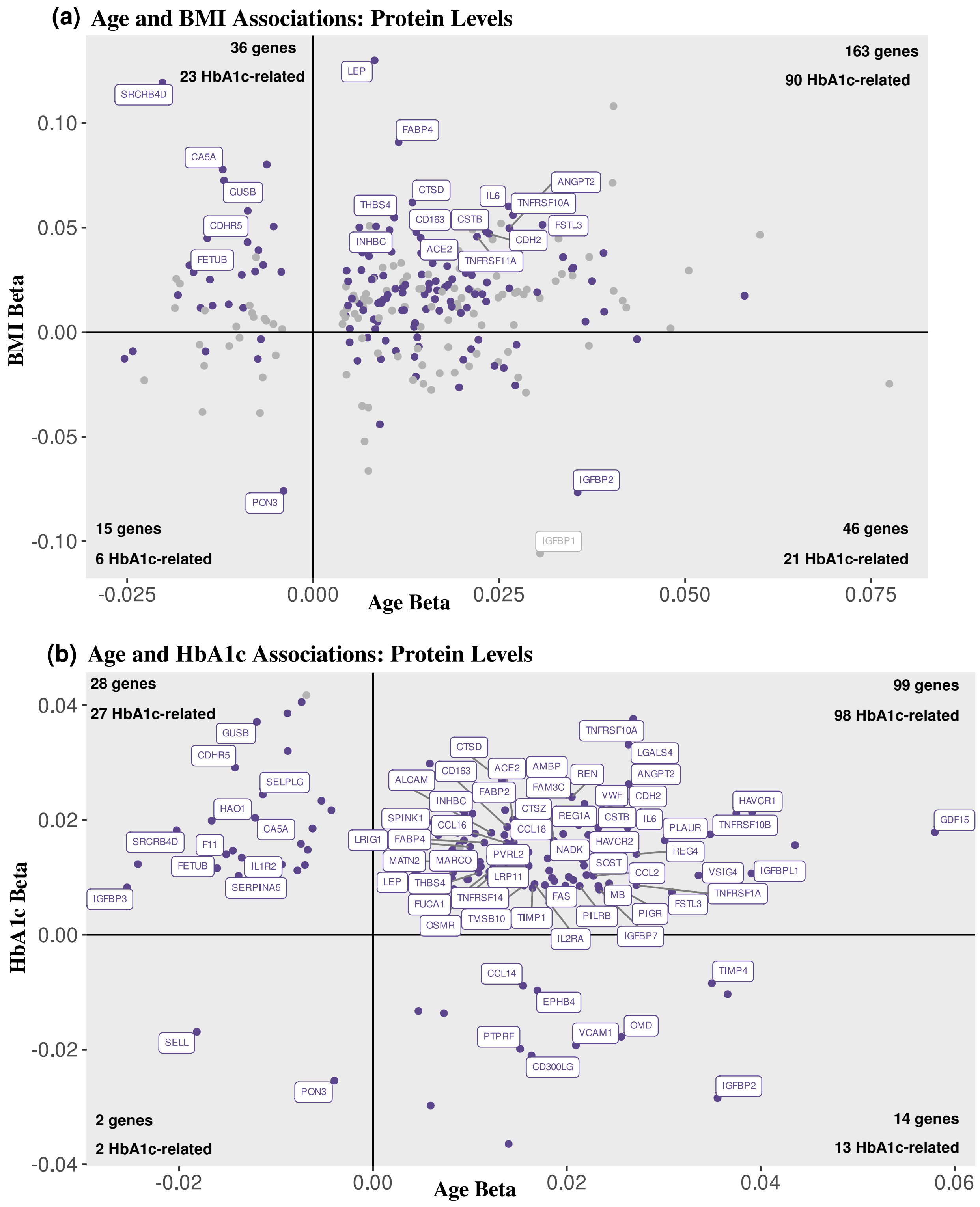


Sup. Figure 17: (a) Proteins independently associated with age and BMI (FDR≤0.05). Proteins with a positive age beta (right side of y-axis) showed increased levels with age while proteins with a negative beta (left side of y-axis) showed decreased levels with age. Similarly, proteins with a positive BMI beta (above x-axis) showed increased levels for higher BMI while proteins with a negative BMI beta (below x-axis) showed decreased levels for higher BMI values. Proteins in purple were also significantly associated with HbA1c. (b) Proteins independently associated with age and HbA1c (FDR≤0.05). Proteins with a positive age beta (right side of y-axis) showed increased levels with age while proteins with a negative beta (left side of y-axis) showed decreased levels with age. Similarly, proteins with a positive HbA1c beta (above x-axis) showed increased levels for higher HbA1c while proteins with a negative HbA1c beta (below x-axis) showed decreased levels for higher HbA1c values. Proteins in purple were also significantly associated with BMI.

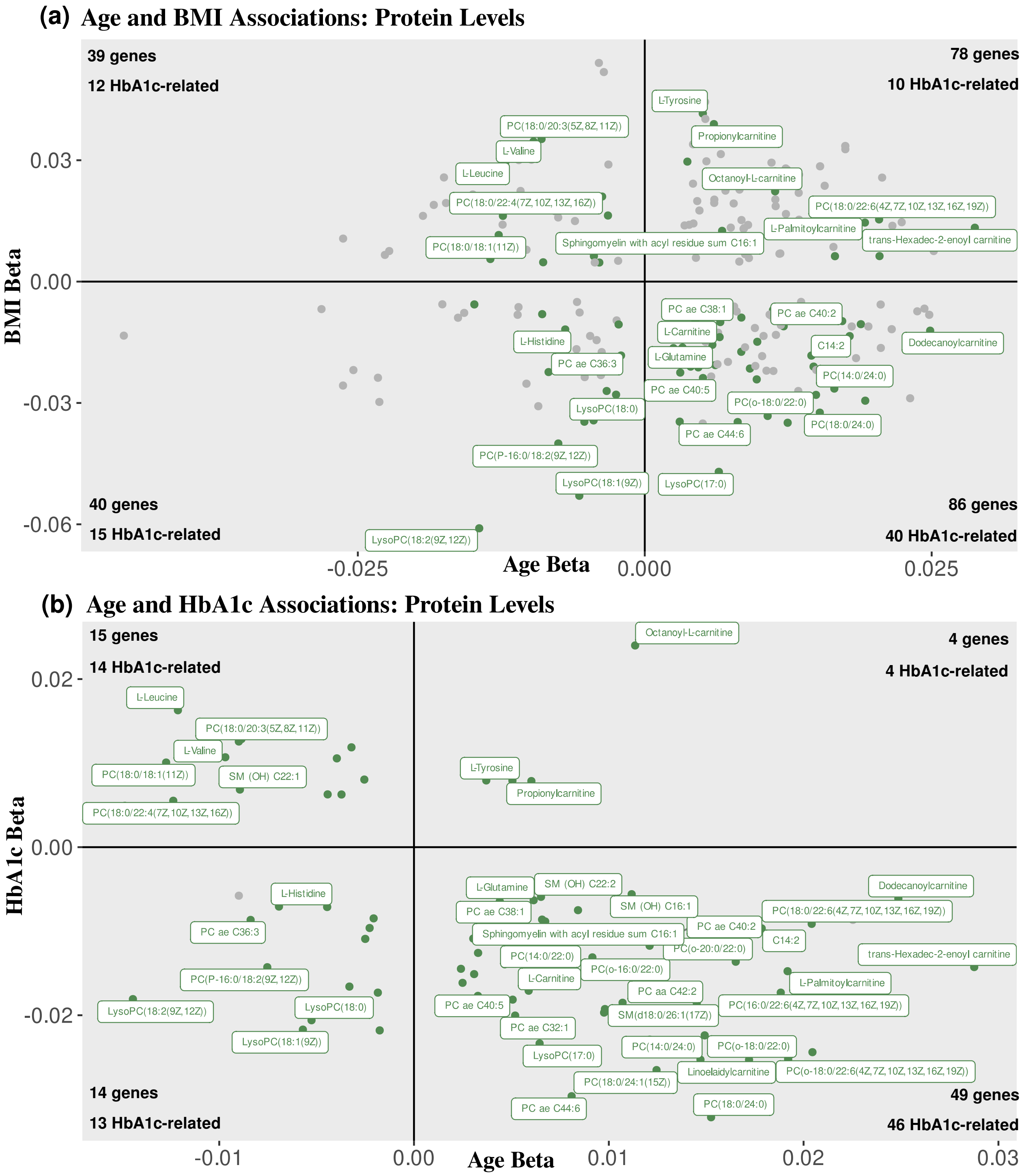


Sup. Figure 18: (a) Metabolites independently associated with age and BMI (FDR≤0.05). Metabolites with a positive age beta (right side of y-axis) showed increased levels with age while metabolites with a negative beta (left side of y-axis) showed decreased levels with age. Similarly, metabolites with a positive BMI beta (above x-axis) showed increased levels for higher BMI while metabolites with a negative BMI beta (below x-axis) showed decreased levels for higher BMI values. Metabolites in green were also significantly associated with HbA1c. (b) Metabolites independently associated with age and BMI (FDR≤0.05). Metabolites with a positive age beta (right side of y-axis) showed increased levels with age while metabolites with a negative beta (left side of y-axis) showed decreased levels with age. Similarly, metabolites with a positive HbA1c beta (above x-axis) showed increased levels for higher HbA1c while metabolites with a negative HbA1c beta (below x-axis) showed decreased levels for higher HbA1c values. Metabolites in green were also significantly associated with BMI.

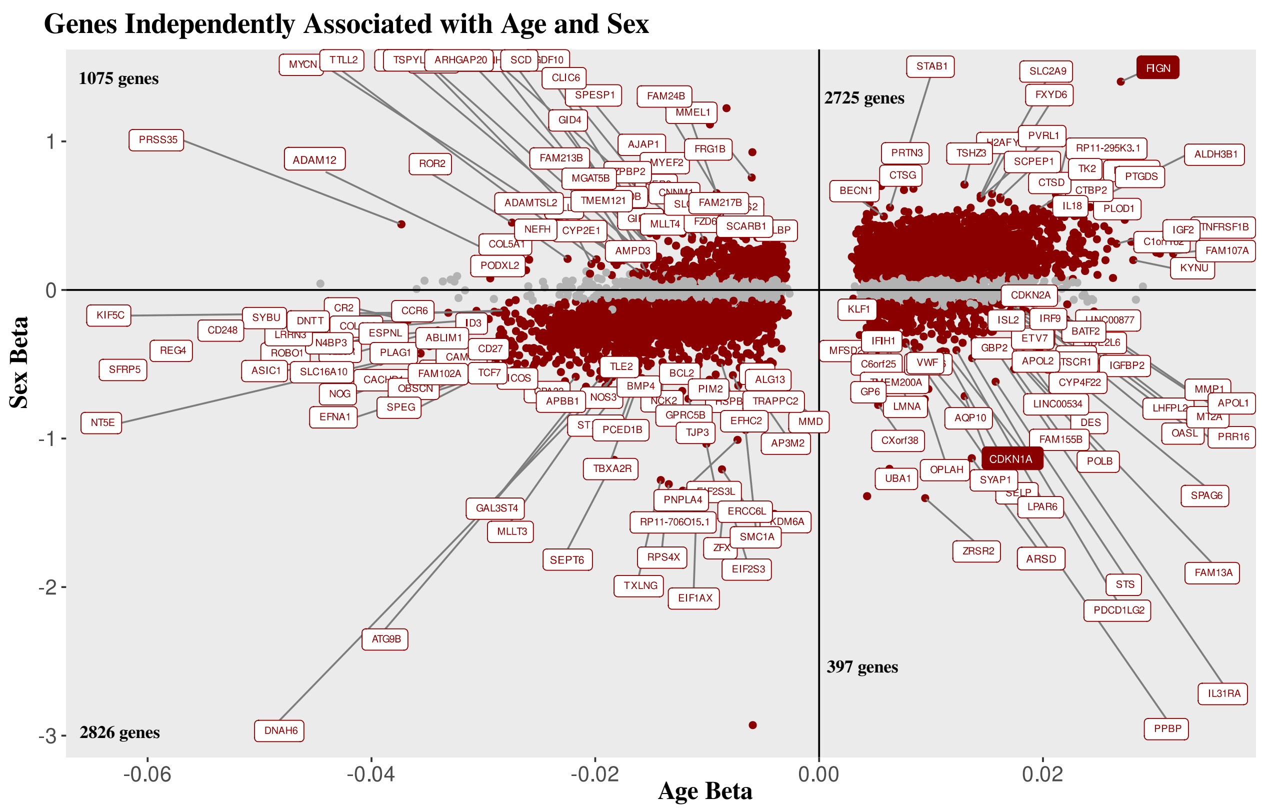


Sup. Figure 19: Genes independently associated with age and sex (FDR≤0.05). Genes with a positive age beta (right side of y-axis) increase with age while genes with a negative beta (left side of y-axis) decrease with age. Similarly, genes with a positive sex beta (above x-axis) have a higher expression in male individuals while genes with a negative sex beta (below x-axis) have a lower expression in male compared to female individuals.

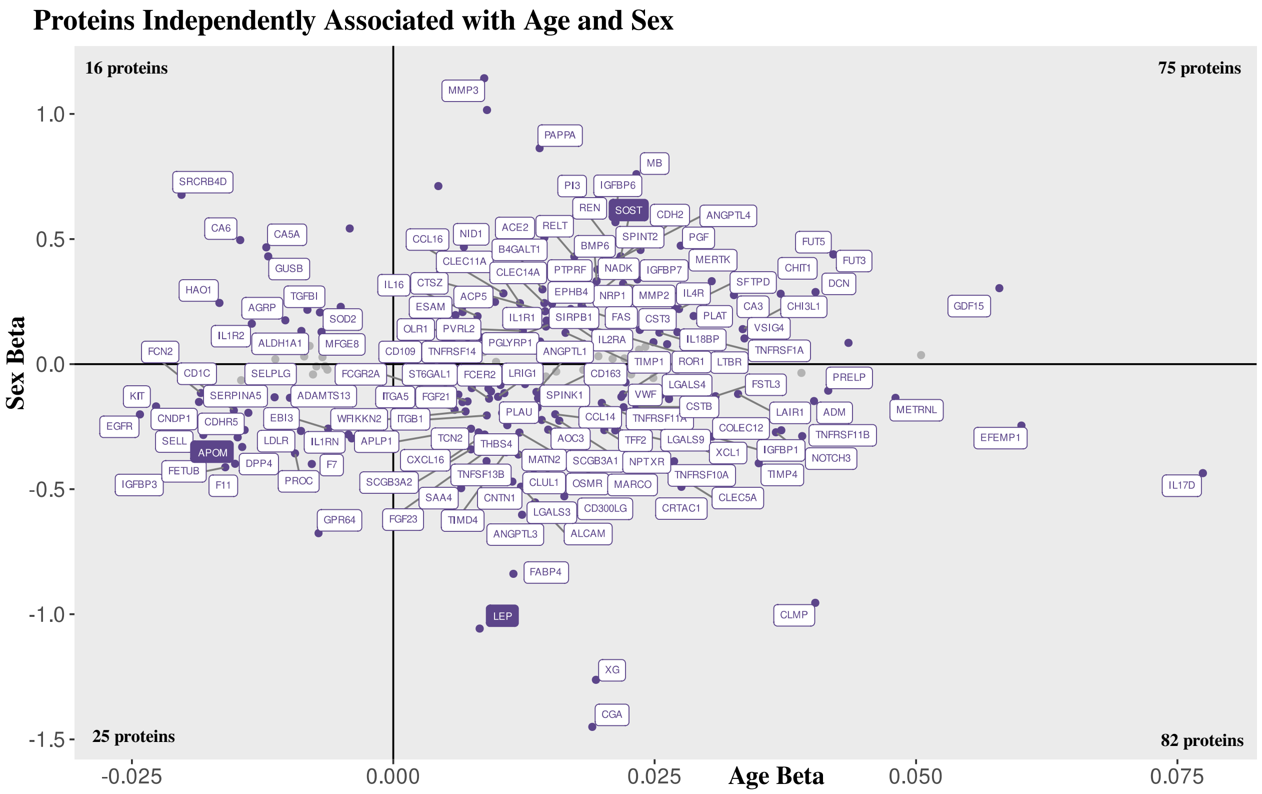


Sup. Figure 20: Proteins independently associated with age and sex (FDR≤0.05). Proteins with a positive age beta (right side of y-axis) increase with age while proteins with a negative beta (left side of y-axis) decrease with age. Similarly, proteins with a positive sex beta (above x-axis) have higher levels in male individuals while proteins with a negative sex beta (below x-axis) have lower levels in male compared to female individuals.

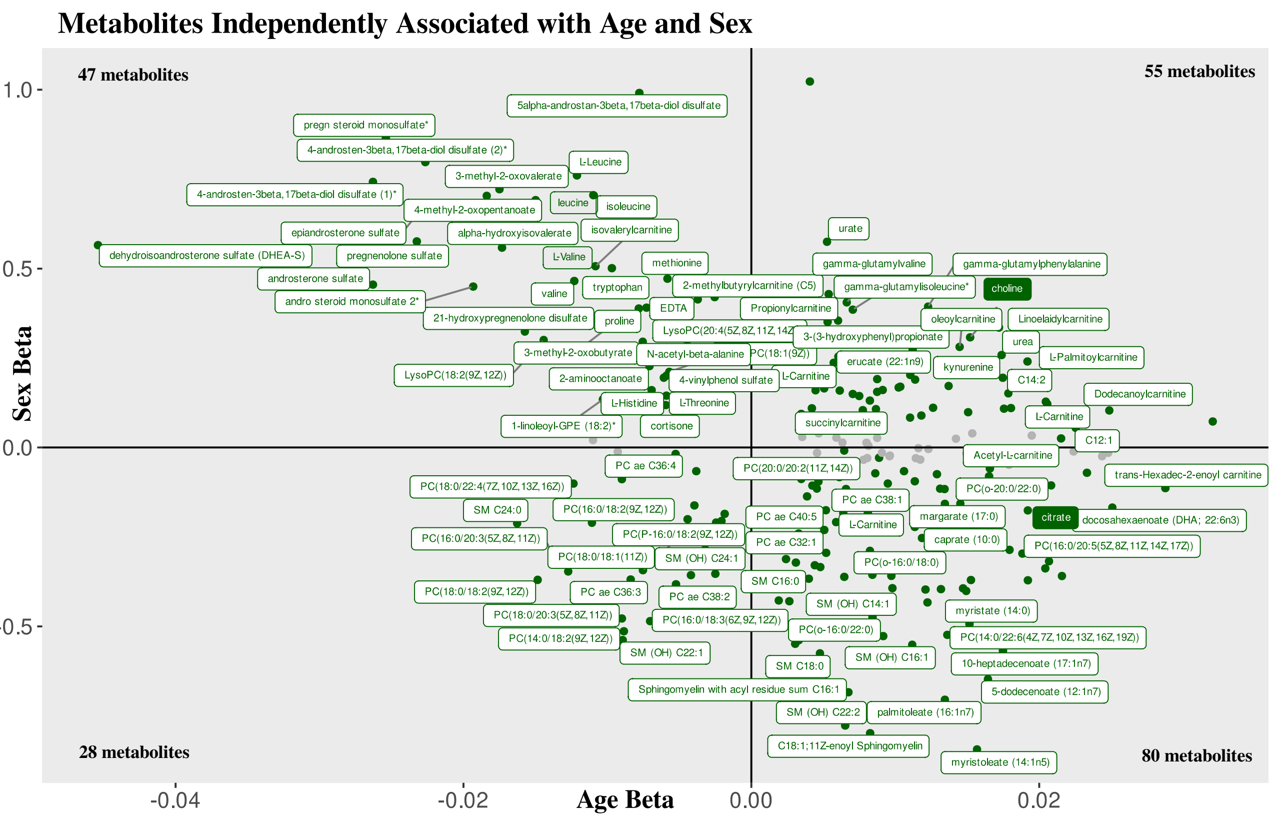


Sup. Figure 21: Metabolites independently associated with age and sex (FDR≤0.05). Metabolites with a positive age beta (right side of y-axis) increase with age while metabolites with a negative beta (left side of y-axis) decrease with age. Similarly, metabolites with a positive sex beta (above x-axis) have higher levels in male individuals while metabolites with a negative sex beta (below x-axis) have lower levels in male compared to female individuals.

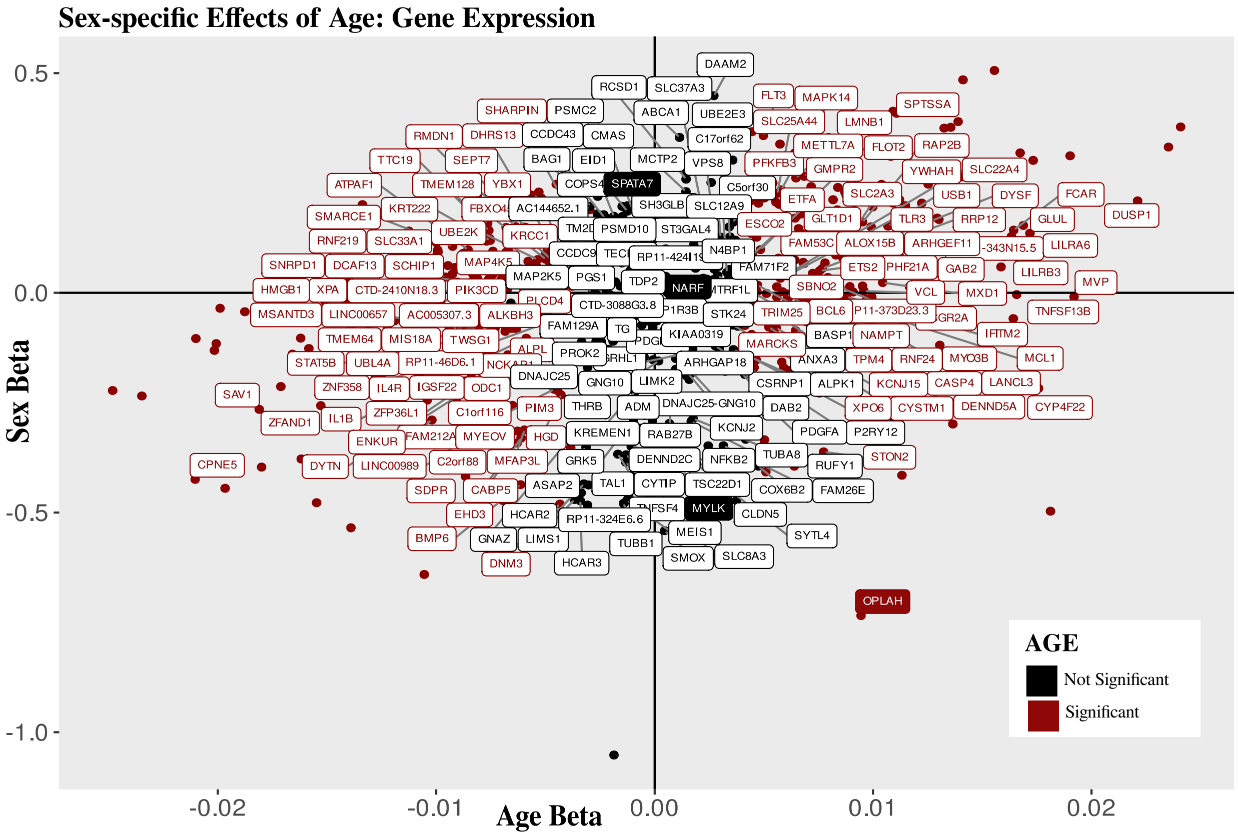


Sup. Figure 22: Genes undergoing sex-specific age effects (age-by-sex interaction, FDR≤0.05). Genes in red were also significantly associated with age (FDR≤0.05) when considering all individuals in the cohort. Expression for genes on the right side of the y-axis collectively increased with age while for genes on the left side of the y-axis collectively decreased with age.

**Sex-specific Effects of Age: Proteins**

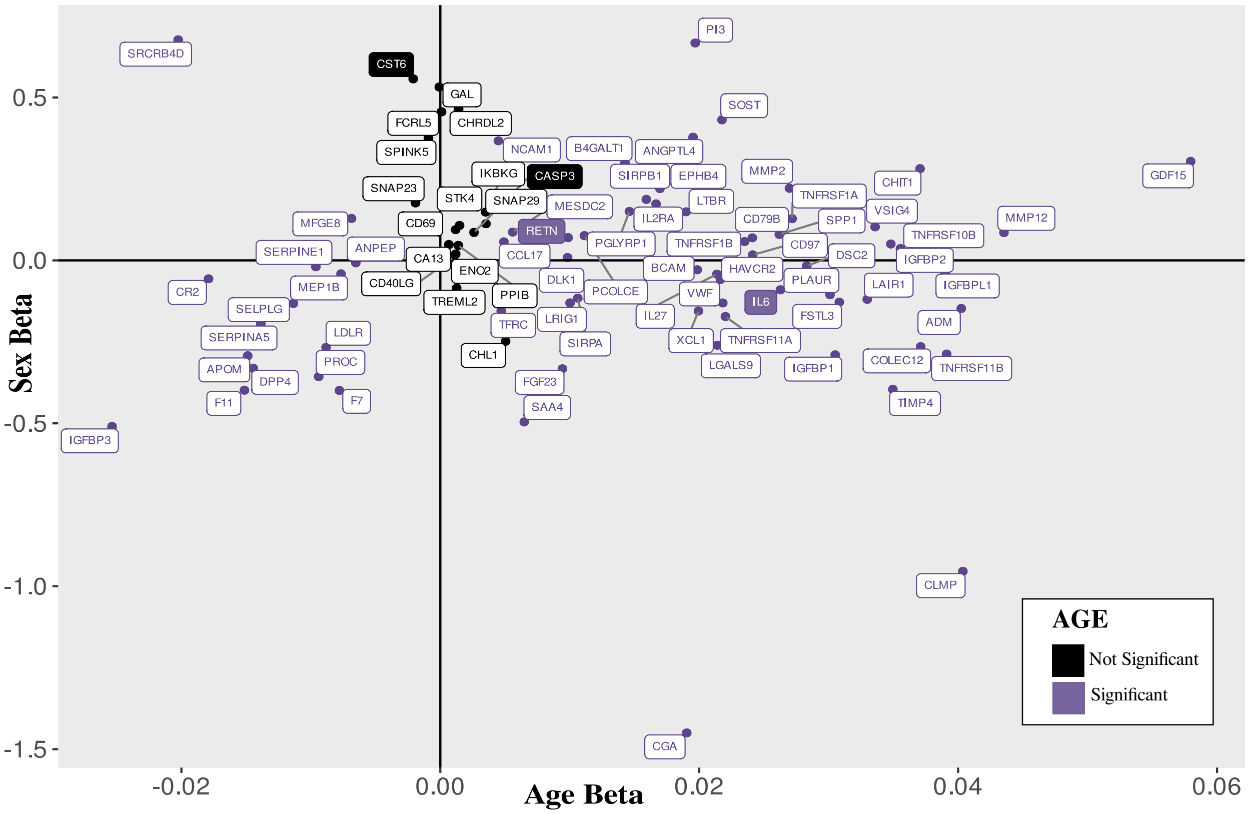


Sup. Figure 23: Proteins undergoing sex-specific age effects (age-by-sex interaction, FDR≤0.05). Proteins in purple were also significantly associated with age (FDR≤0.05) when considering all individuals in the cohort. Levels for proteins on the right side of the y-axis collectively increased with age while for proteins on the left side of the y-axis collectively decreased with age.

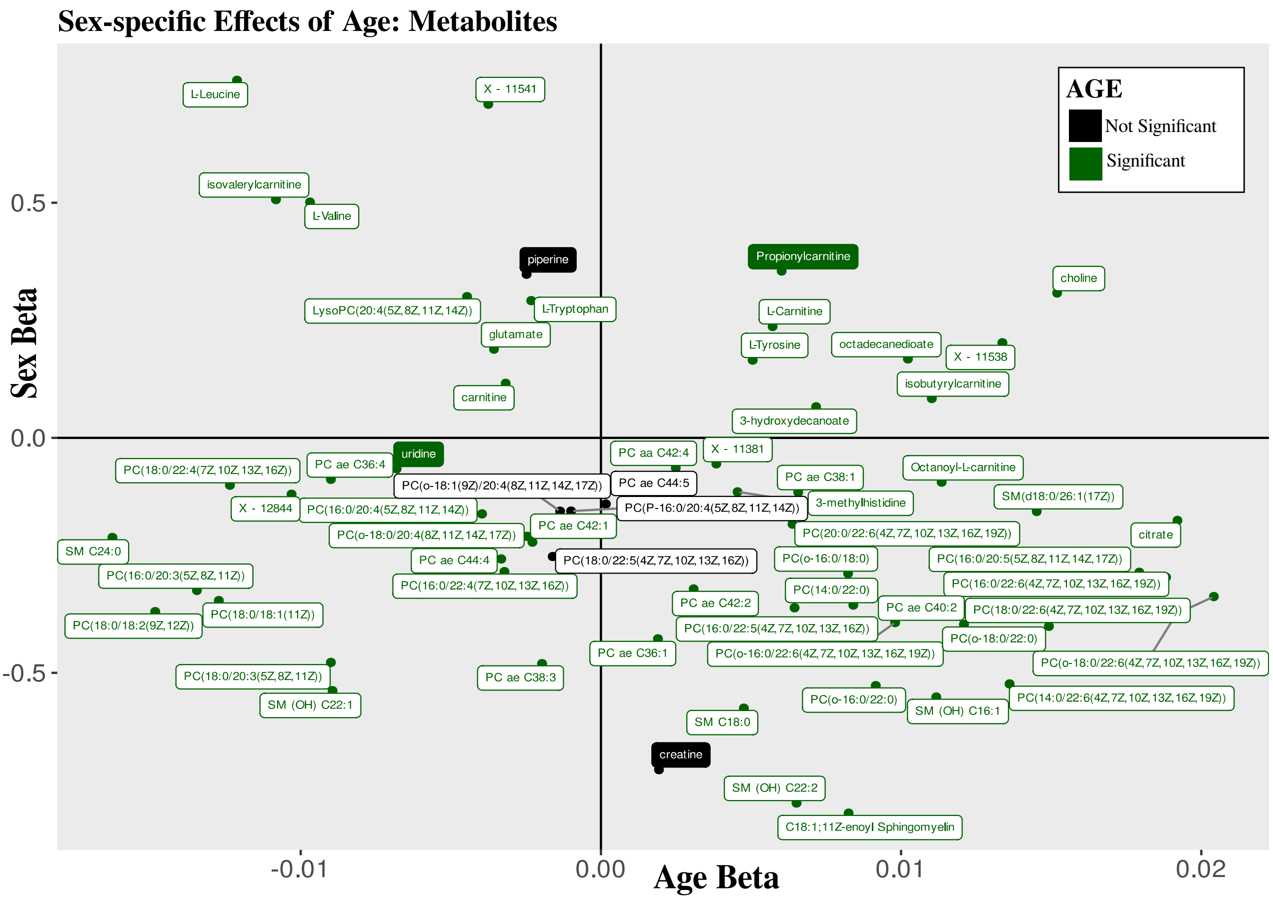


Sup. Figure 24: Metabolites undergoing sex-specific age effects (age-by-sex interaction, FDR≤0.05). Metabolites in green were also significantly associated with age (FDR≤0.05) when considering all individuals in the cohort. Levels for metabolites on the right side of the y-axis collectively increased with age while for metabolites on the left side of the y-axis collectively decreased with age.

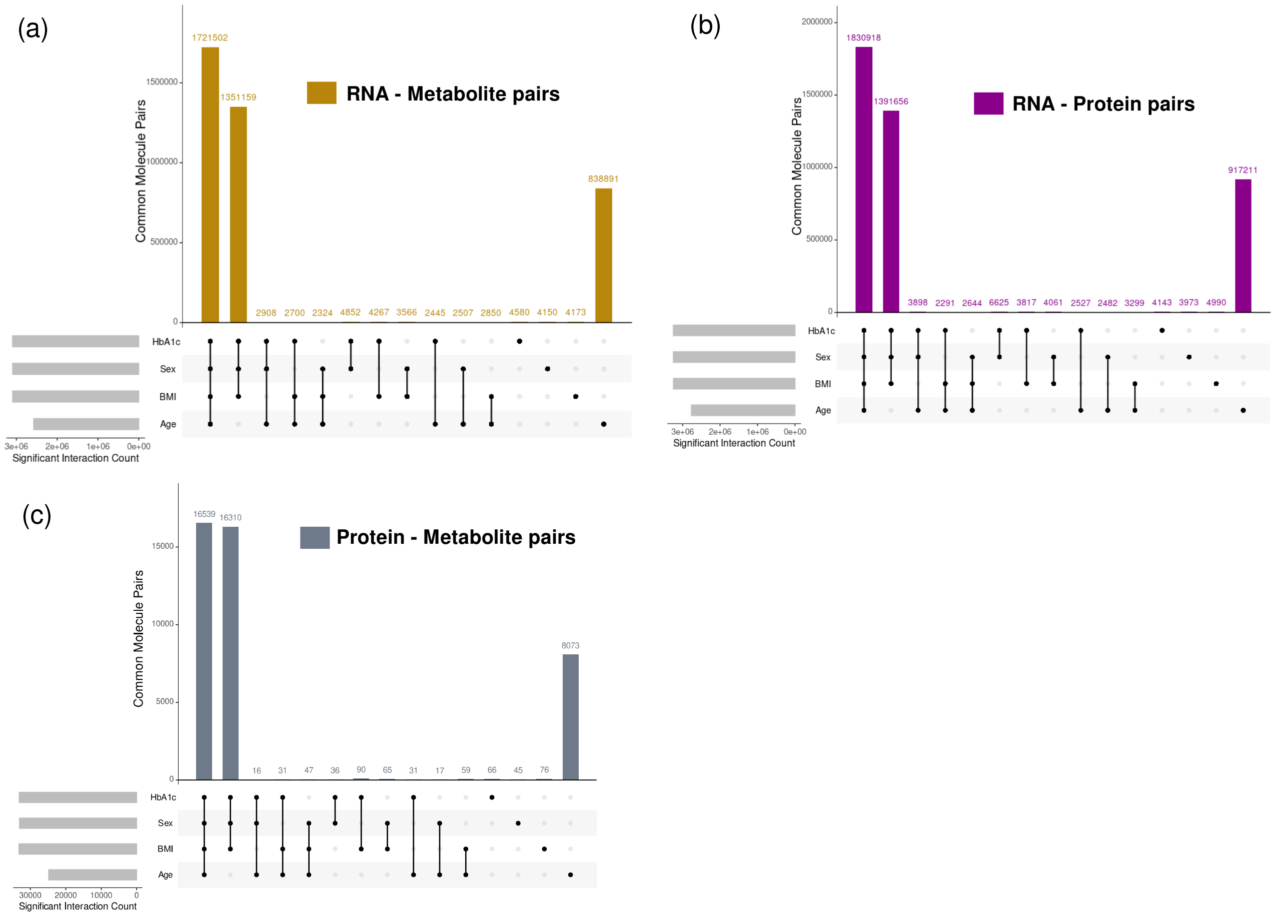


Sup. Figure 25: Upset plots showing all intersections of molecule pair relationships modulated by all biological traits tested: (a) Overlap of RNA-metabolite relationships influenced by HbA1c, sex, BMI and age. (b) Overlap of RNA-protein relationships influenced by HbA1c, sex, BMI and age. (c) Protein-metabolite relationships influenced by HbA1c, sex, BMI and age.

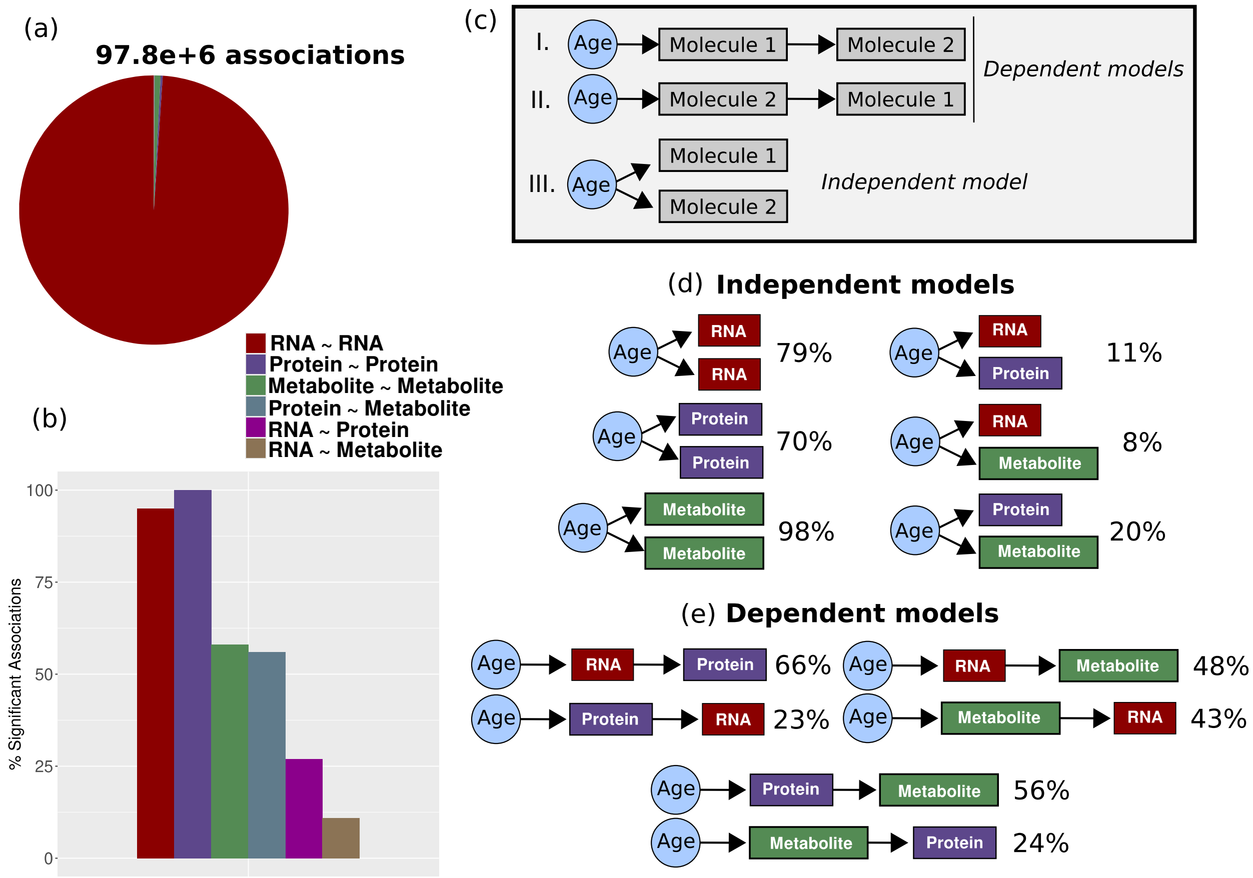


Sup. Figure 26: Direction of causality in age-related molecular phenotype relationships: (a) Relative counts of significantly associated age-related molecule pairs per comparison set between molecule types; we identified 97,807,420 associations (84% of tested pairs) the majority of which involved association between gene expression. (b) Percentages of significantly associated age-related molecules pairs per comparison set between molecule types. (c) Three models were tested for casual inference in order to evaluate the direction of causality between age and molecular phenotype pairs. The dependent models assume that the effect of age on one molecule (1 or 2) is mediated by the other molecule and vice versa. The independent model assumes that the effect of age on both molecular phenotypes is independent, thus there is no association/mediation present between the two molecules. (d) Percentages of relationships more likely to be explained by the independent model according to the causal inference analysis results. (e) Percentages of relationships more likely to be explained by a dependent model according to the causal inference analysis results. Percentages were calculated against the number of pairs for which we could get a supported decision (BIC $\geq$ 6; 88,836,252 pairs; 91% of identified molecule associations).

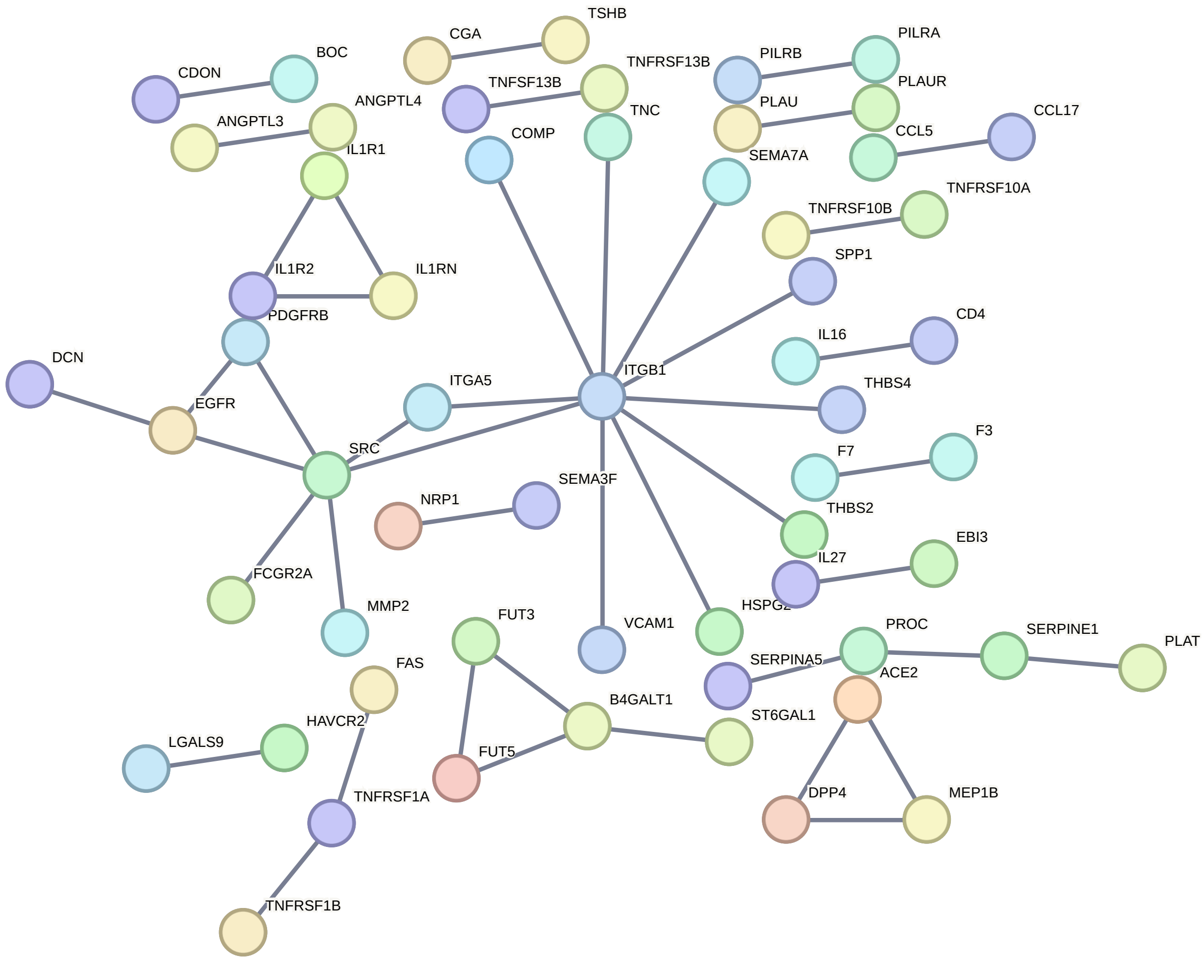


Sup. Figure 27: Protein-Protein Interaction (PPI) network of 267 age-related proteins. Network was constructed using STRING database, extracting interaction information from databases and plotting only connections between proteins (nodes) with the highest confidence per interaction (0.9).

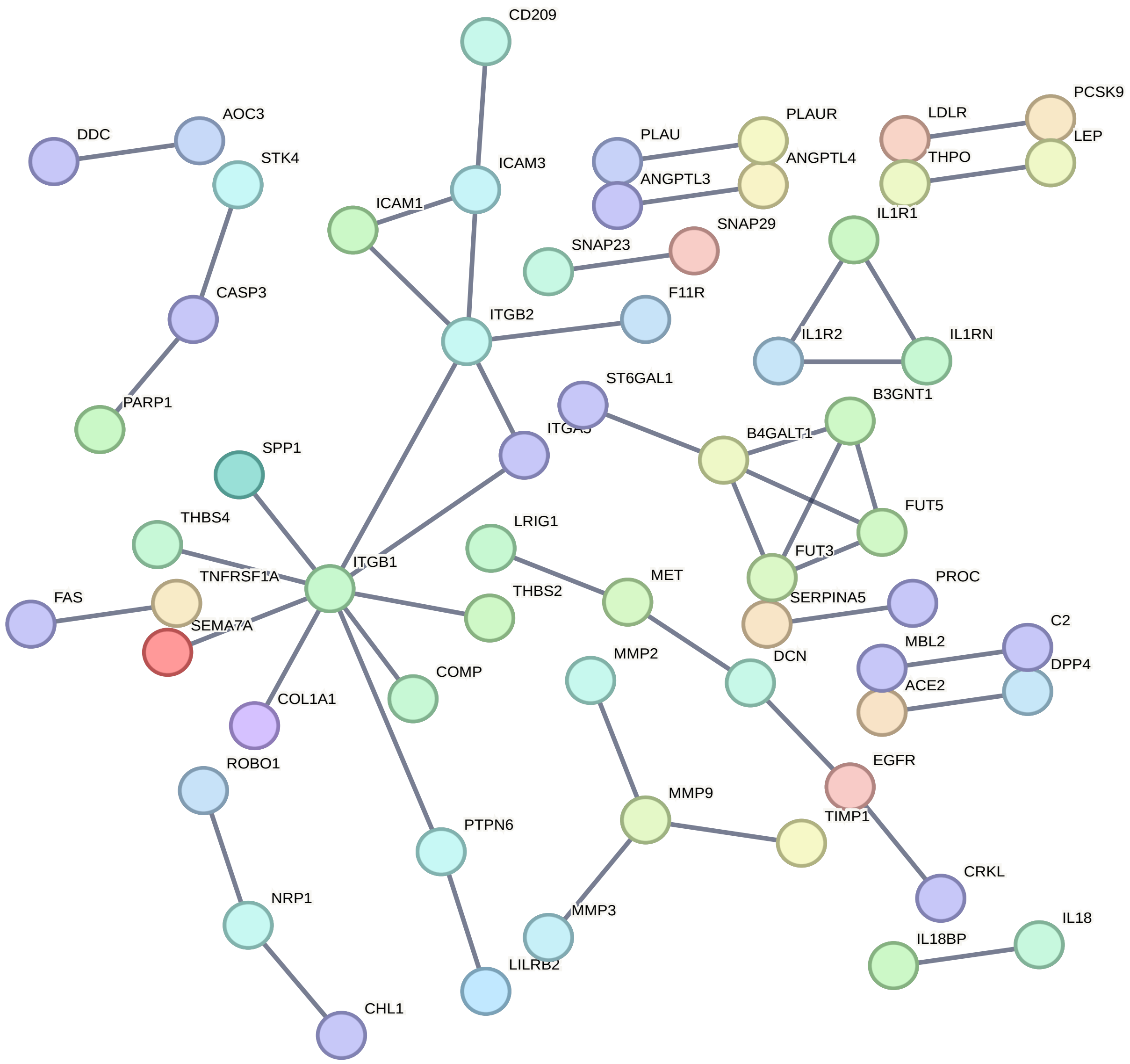


Sup. Figure 28: Protein-Protein Interaction (PPI) network of 267 sex-related proteins. Network was constructed using STRING database, extracting interaction information from databases and plotting only connections between proteins (nodes) with the highest confidence per interaction (0.9).

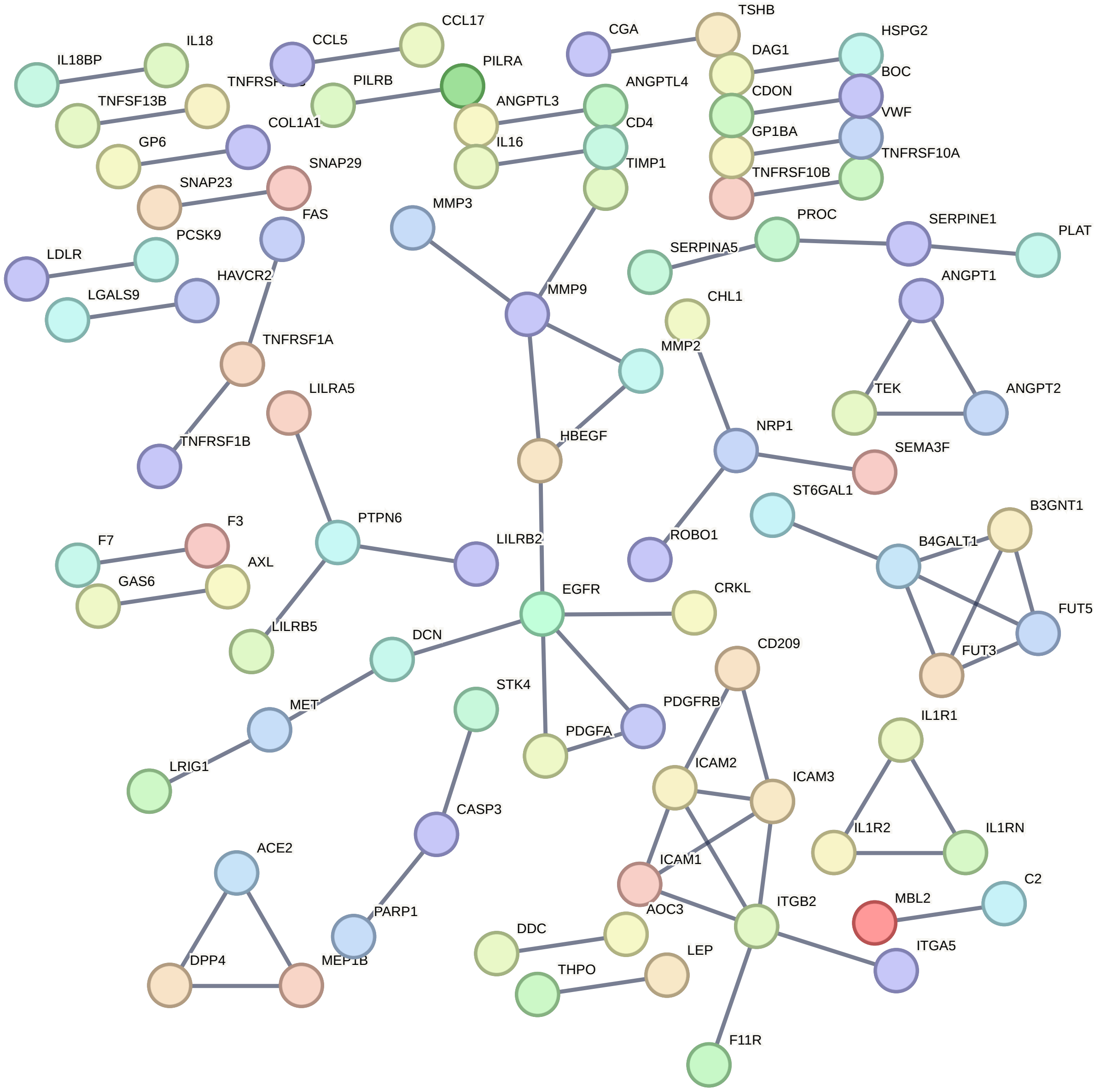


Sup. Figure 29: Protein-Protein Interaction (PPI) network of 363 BMI-related proteins. Network was constructed using STRING database, extracting interaction information from databases and plotting only connections between proteins (nodes) with the highest confidence per interaction (0.9).

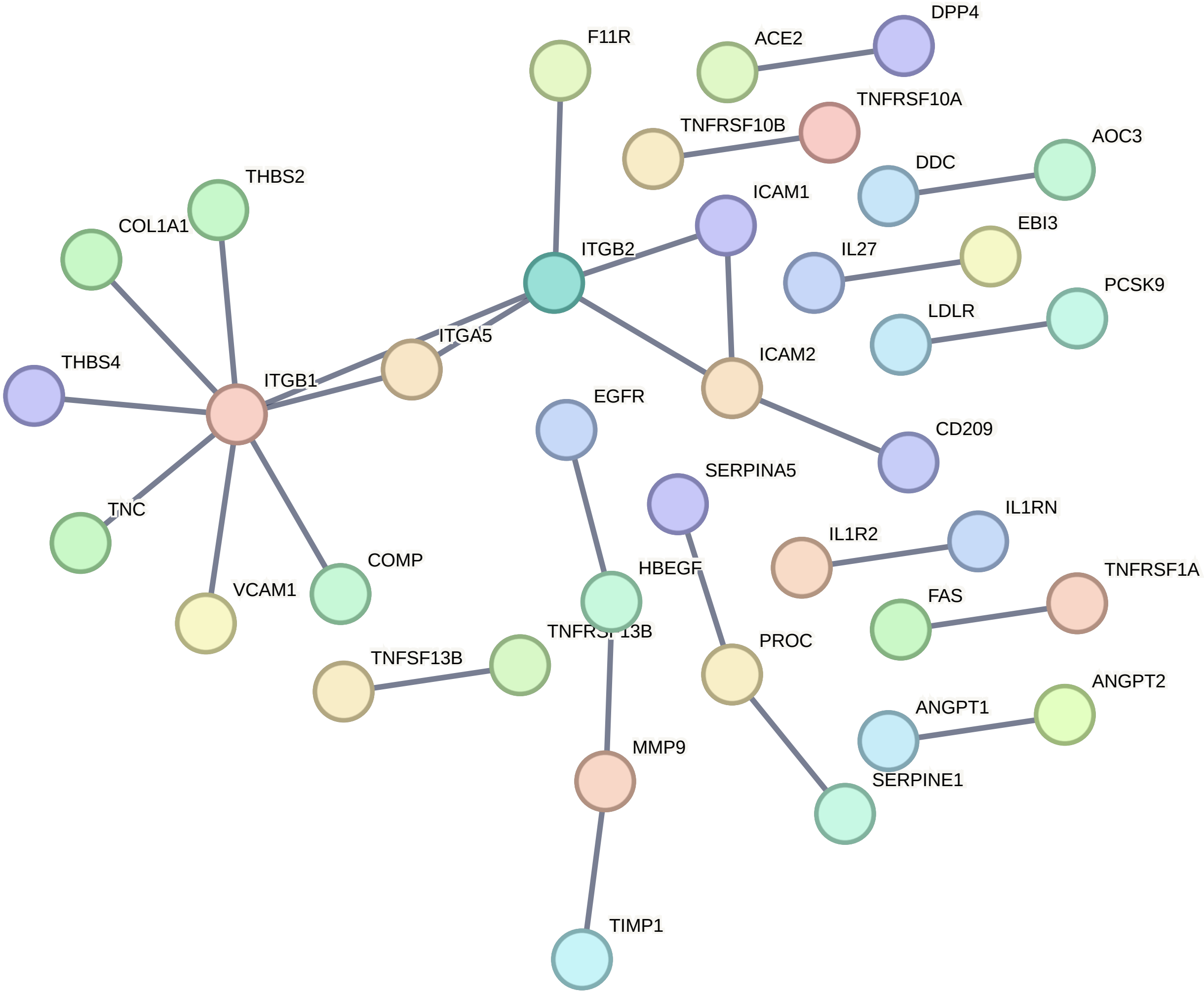


Sup. Figure 30: Protein-Protein Interaction (PPI) network of 193 HbA1c-related proteins. Network was constructed using STRING database, extracting interaction information from databases and plotting only connections between proteins (nodes) with the highest confidence per interaction (0.9).
